# Speech and language impairments in behavioral variant frontotemporal dementia: A systematic review

**DOI:** 10.1101/2021.07.10.21260313

**Authors:** Amandine Geraudie, Petronilla Battista, Adolfo M. García, Isabel E. Allen, Zachary A. Miller, Maria Luisa Gorno-Tempini, Maxime Montembeault

## Abstract

Although behavioral variant frontotemporal dementia (bvFTD) is classically defined by behavioral and socio-emotional changes, impairments often extend to other cognitive functions. These include early speech and language deficits related to the disease’s core neural disruptions. Yet, their scope and clinical relevance remains poorly understood. This systematic review characterizes such disturbances in bvFTD, considering clinically, neuroanatomically, genetically, and neuropathologically defined subgroups. We included 181 experimental studies, with at least 5 bvFTD patients diagnosed using accepted criteria, comparing speech and language outcomes between bvFTD patients and healthy controls or between bvFTD subgroups. Results reveal extensive and heterogeneous deficits across cohorts, with (a) consistent lexico-semantic, orthographic, and prosodic impairments; (b) inconsistent deficits in motor speech and grammar; and (c) relative preservation of phonological skills. Also, preliminary findings suggest that the severity of speech and language deficits might be associated with global cognitive impairment, predominantly temporal or fronto-temporal atrophy and *MAPT* mutations (vs *C9orf72*). Although under-recognized, these impairments contribute to patient characterization and phenotyping, while potentially informing diagnosis and prognosis.

## 1. Introduction

The behavioral variant of frontotemporal dementia (bvFTD) is a neurodegenerative disorder characterized by a progressive decline in social cognitive functions and changes in personality and behavior (Neary et al., 1998; Rascovsky et al., 2011). Current diagnostic criteria include early behavioral disinhibition, apathy or inertia, loss of sympathy or empathy, stereotyped behaviour, hyperorality, dietary changes, deficits in executive functions and spared memory and visuo-spatial functions (Rascovsky et al., 2011). Yet, substantial deficits are being increasingly recognized in other domains. Crucially, these include speech and language deficits, which often emerge in early disease stages (Cheran et al., 2019) and affect daily functionality (Lima-Silva et al., 2015). To understand the scope and clinical relevance of these disruptions, we conducted the first systematic review of speech and language deficits in bvFTD, considering their relationship with clinical, neural, genetic, and neuropathological factors.

Beyond a brief mention of stereotypy of speech, current diagnostic criteria of bvFTD make no reference to speech and language. Yet, the article describing the criteria report specific speech and language deficits in as many 20% of patients (Rascovsky et al., 2011), in line with earlier characterizations highlighting altered speech output (aspontaneity or press of speech), stereotypies, echolalia, perseveration, and mutism (Neary et al., 1998). More recently, evidence from a large bvFTD sample (Saxon et al., 2017) shows that naming deficits are as frequent as hyperorality (a core diagnostic feature) in the sample informing Rascovsky’s criteria (55%) (Rascovsky et al., 2011). In bvFTD, speech and language symptoms extend to multiple language domains and go beyond executive dysfunction (Hardy et al., 2016). They can appear very early in the disease, even at pre-symptomatic stages (Cheran et al., 2019), and they worsen as disease progresses (Ash et al., 2019). They are related to damage in cerebral regions that are targeted in bvFTD as well as part of the speech and language brain networks (Hardy et al., 2016). They can also pose differential diagnosis challenges with language-predominant neurodegenerative diseases such as primary progressive aphasias (PPA) (Pozueta et al., 2019). Thus, the neglect of speech and language in bvFTD may hinder valuable opportunities for disease characterization and phenotyping, calling for comprehensive overviews of existing evidence.

Yet, no study has provided a comprehensive and systematic overview of speech and language in bvFTD. Although a few primary studies have focused on language symptoms in bvFTD, they most often include low sample sizes, given the low prevalence of this syndrome, or they only provide partial coverage of relevant language domains. Fortunately, multiple bvFTD reports also include language tests as part of their secondary outcome measures. A systematic review of all this evidence would provide a detailed account of speech and language in bvFTD, potentially revealing underexplored markers to complement diagnosis (especially in contexts lacking gold-standard pathological or genetic data), prognosis (as communication difficulties may influence behavioural changes (Harris et al., 2016)), and management (given that such difficulties increase caregiver burden (Savundranayagam et al., 2005).

The goal of this study is to systematically review the existing studies covering language functioning in bvFTD. First, we aim to determine the impaired and preserved language domains in bvFTD by reviewing studies comparing bvFTD patients to controls. Given the fact that we expect some heterogeneity in language outcomes in bvFTD patients, the second aim of this study is to investigate which bvFTD features are associated with more severe language impairments. To do so, we will review studies comparing patients’ subgroups based on clinical (for example, according to severity of the global cognitive impairment), neural (for example, temporal vs. frontal atrophy), genetic (for example, no mutation, *C9orf72, MAPT* or *GRN* mutations), and neuropathological (for example, frontotemporal lobar degeneration tau [FTLD-tau] vs. FTLD-TDP) profiles. A better characterization of language profile in these patients may contribute to a more fine-grained phenotyping, differential diagnosis as well as prognosis in bvFTD patients.

## 2. Methods

This work was performed and reported following Preferred Reporting Items for Systematic Reviews and Meta-Analyses (PRISMA) guidelines (Page et al., 2021).

### 2.1. Search strategy

A systematic review of the literature was conducted on the following databases: PubMed, Embase, Web of Science, PsycINFO and Cochrane CENTRAL, with the help of a university librarian. In PubMed, the search was conducted with the following search terms: (“language disorders” (as a MeSH term), “Aphasia” (as a MeSH term), “language”, “linguistic”, “aphasia”, “aphasic”, “speech”, “naming”, “anomic”, “anomia”, “semantic”, “semantics”, “syntax”, “syntactic”, “prosody”, “prosodic”, “phonology”, “phonological”, “discourse”, “picture description”, “fluency”, “grammar”, “agrammatism”, “agrammatic”, “writing”, “write”, “reading”, “read”, “comprehension”, “repetition”, “dysarthria”, “spell”, “spelling”, “communication”, “word”, “sentence”, “verb”, “verbal”, “noun”, “lexical”, “lexicon”, “dyslexia”, “dyslexic”, “alexia”, “alexic”) AND (“bvFTD” or (“behavioral” or “behavioural”) and (“frontotemporal lobar degeneration” (as a MeSH term) or “ftld”) or ((“behavioral” or “behavioural”) and (“frontotemporal” or “fronto-temporal”) and (“dementia” or “dementias” or “degeneration”)) or ((“behavioral” or “behavioural”) and (“ftd”)). The same terms were used for a generic search (without PubMed field tags) in other databases. The search was conducted on February 9^th^, 2020. Additionally, reference lists of identified papers were manually reviewed for additional relevant articles. Duplicates were removed and only published articles or articles in press were retained.

### 2.2. Study selection

Articles were selected by two independent raters (AG and MM) based on the following criteria: (i) article written in English; (ii) experimental study in humans (excluding reviews and animal studies); (iii) inclusion of at least five bvFTD patients (excluding FTD-ALS patients or general FTD group); (iv) bvFTD patients diagnosed with accepted international consensus criteria (Neary et al., 1998; Rascovsky et al., 2011) – in light of this criterion, articles published before 1998 have therefore been excluded; when the diagnostic approach was not clear, the corresponding author of the paper was contacted by email (in case of no response or unavailability of relevant information from the researchers, the study was excluded); (v) comparison group of interest (healthy controls or subgroups of bvFTD patients); (vi) report of the performance in a relevant speech or language domain for bvFTD patients alongside a statistical comparison between groups. We targeted the following speech and language domains: global language screening, motor speech (including motor speech examination and motor speech features extracted from connected speech), phonology (including phonological manipulation and repetition tests), orthography (including reading and writing tasks), lexico-semantics (including word retrieval, word comprehension, semantic knowledge and semantic features extracted from connected speech), grammar (including sentence comprehension and grammatical features extracted from connected speech), and prosody (including receptive and productive tasks). To determine inclusion, the two authors (AG, MM) separately reviewed first the titles and abstract and then the full-text papers. For studies in which there was a disagreement, consensus was reached through discussion between both authors.

### 2.3. Data extraction

Our final selection comprised 181 papers (Figure 1), each of which was used to extract the following information: (i) bvFTD sample characteristics (sample size, mean age, mean disease duration, mean severity using Mini-Mental State Exam), (ii) sample size of the comparison group, (iii) language tests administered, (iv) results of statistical comparison for language measures, (v) participant’s language, (vi) research or clinical center in which patients were recruited. Since the same patient cohort may have been used for multiple studies from a single research team, we flagged results on the exact same language measures from the same team and used a systematic approach to ensure exhaustivity and reduce population bias. First, we reported all studies (no matter the research center) in order to give a complete overview of the literature (Table 1). However, in the Results section and in Figure 2, if two or more studies from the same research center included the exact same language task, only the article with the largest sample of bvFTD patients was considered for interpretation.

**Table 1.**
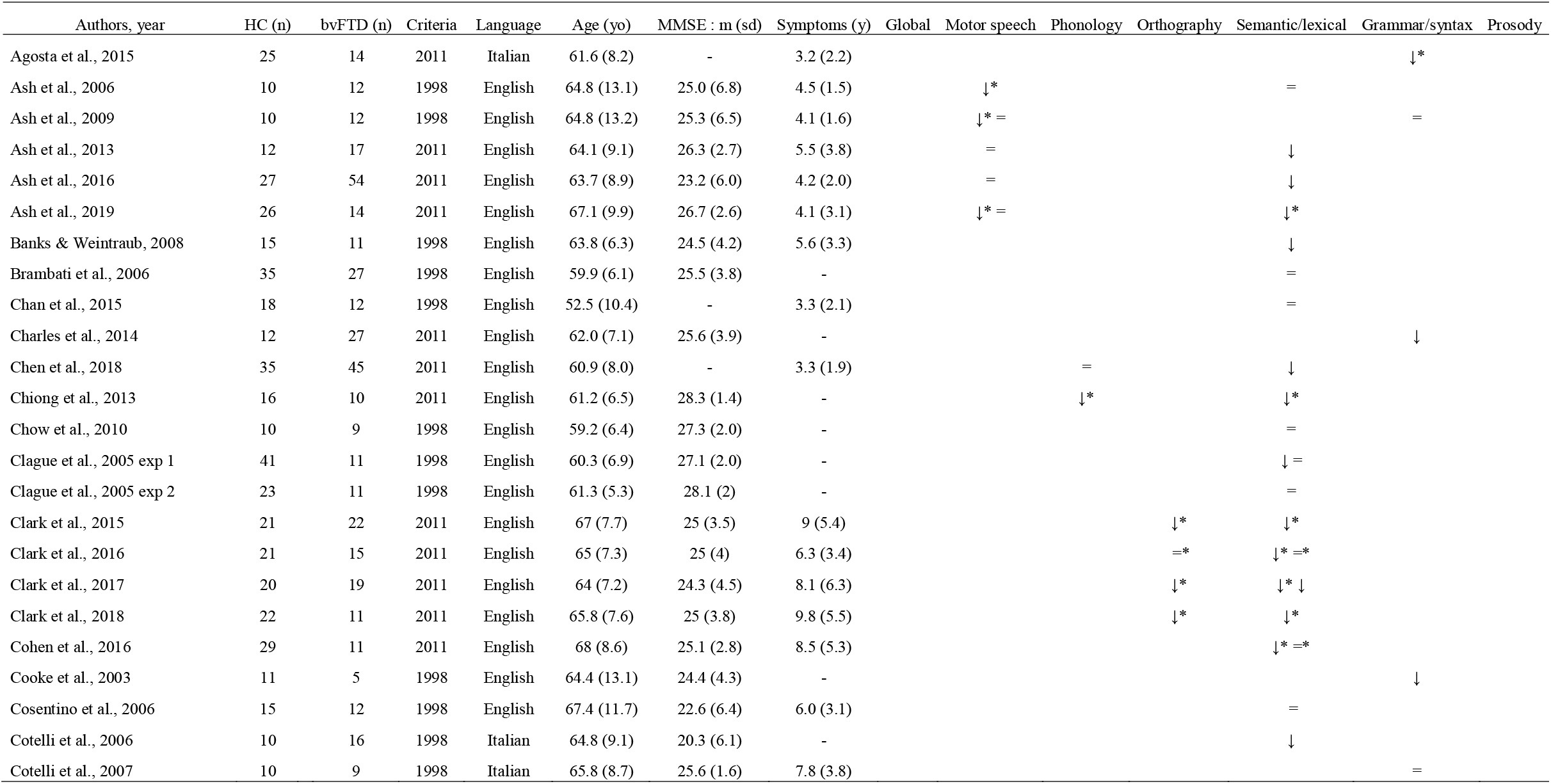
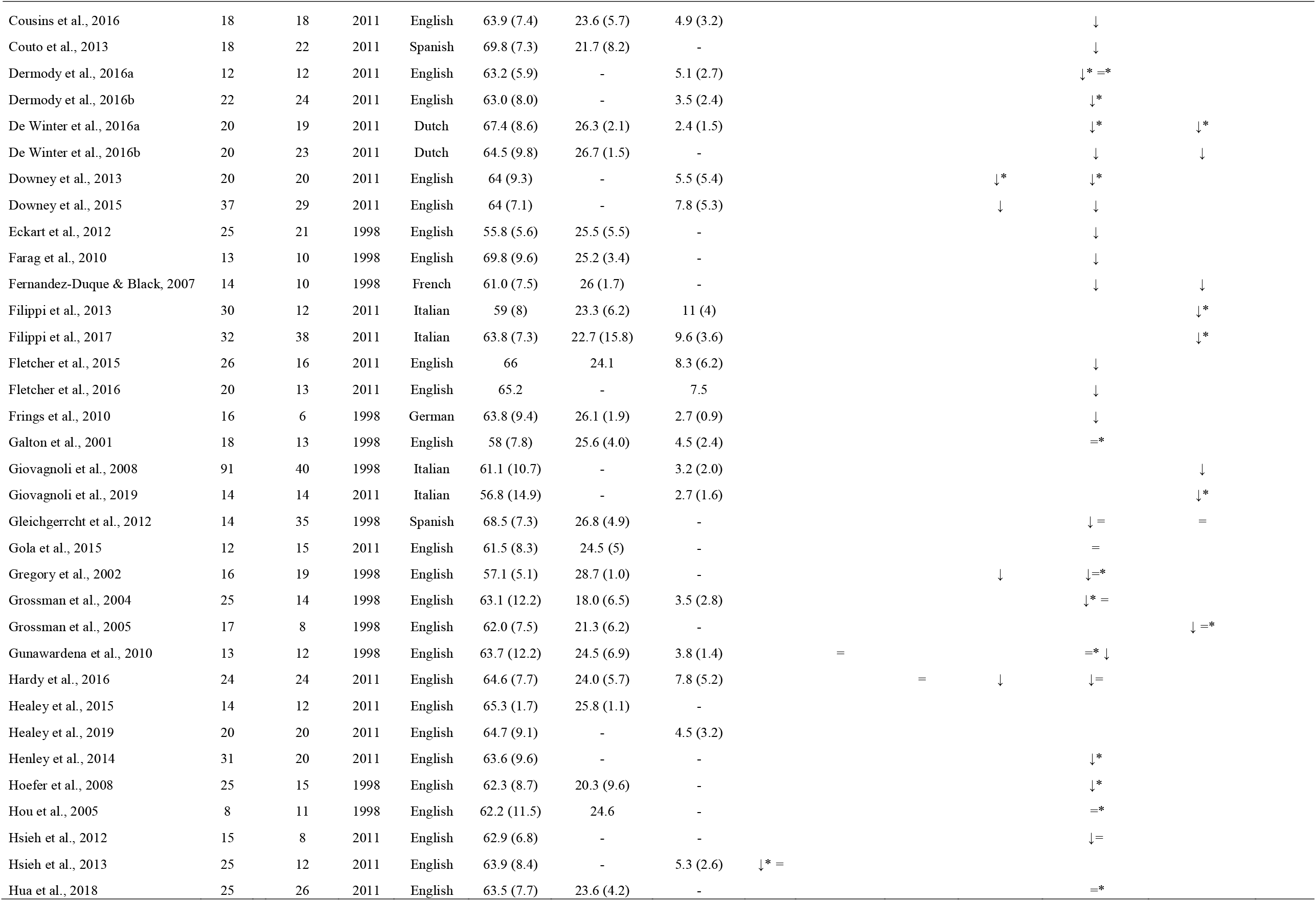
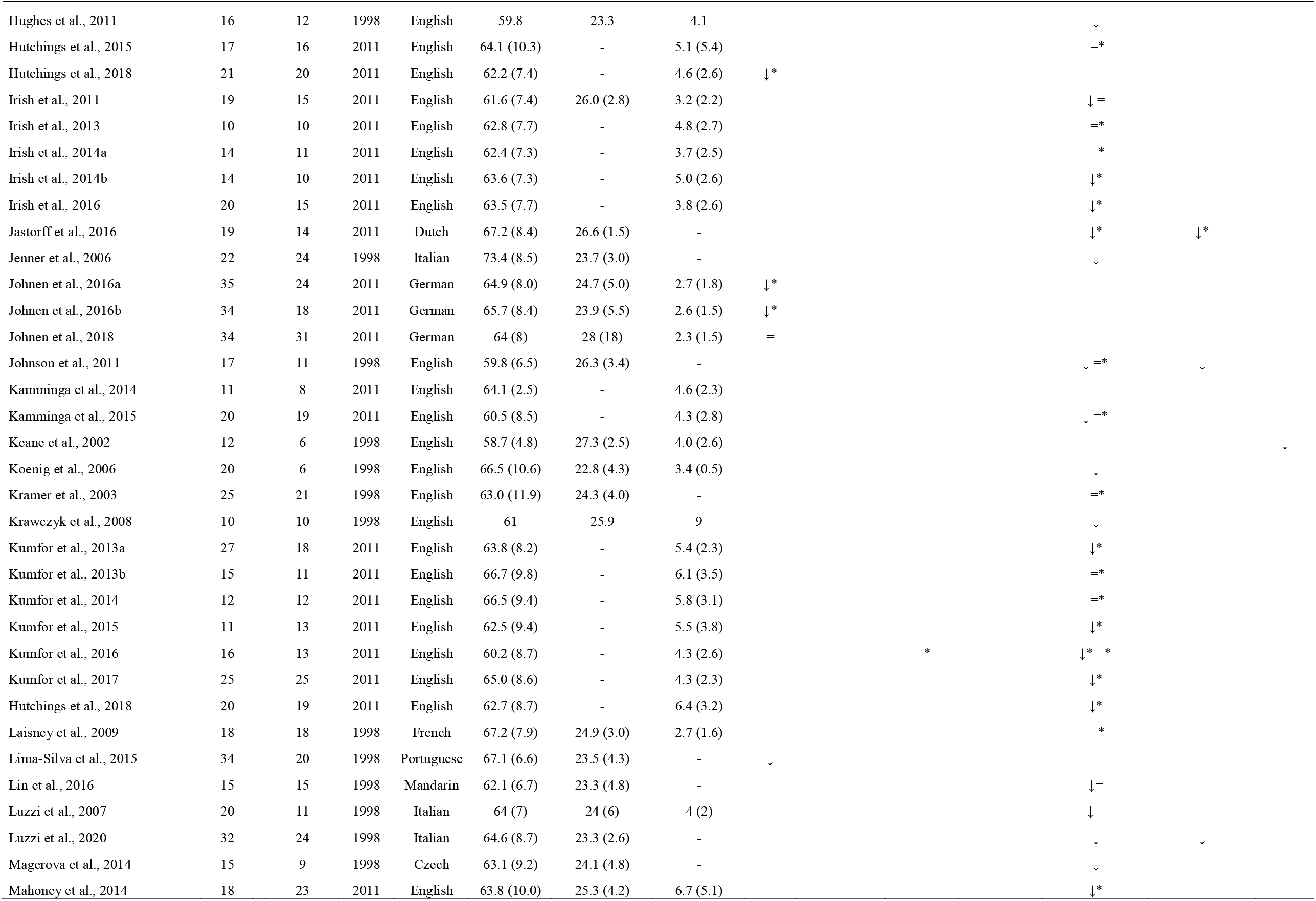
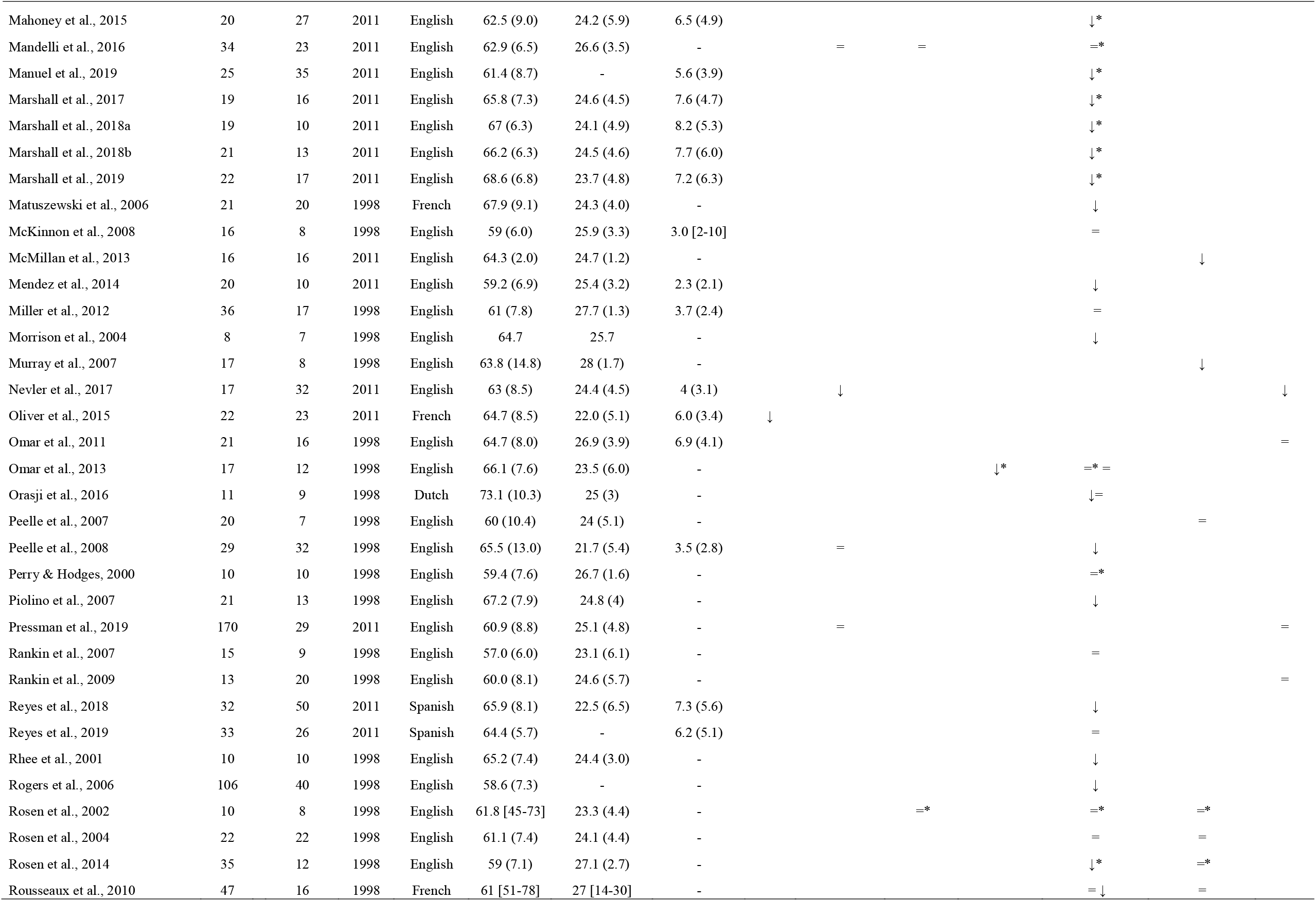
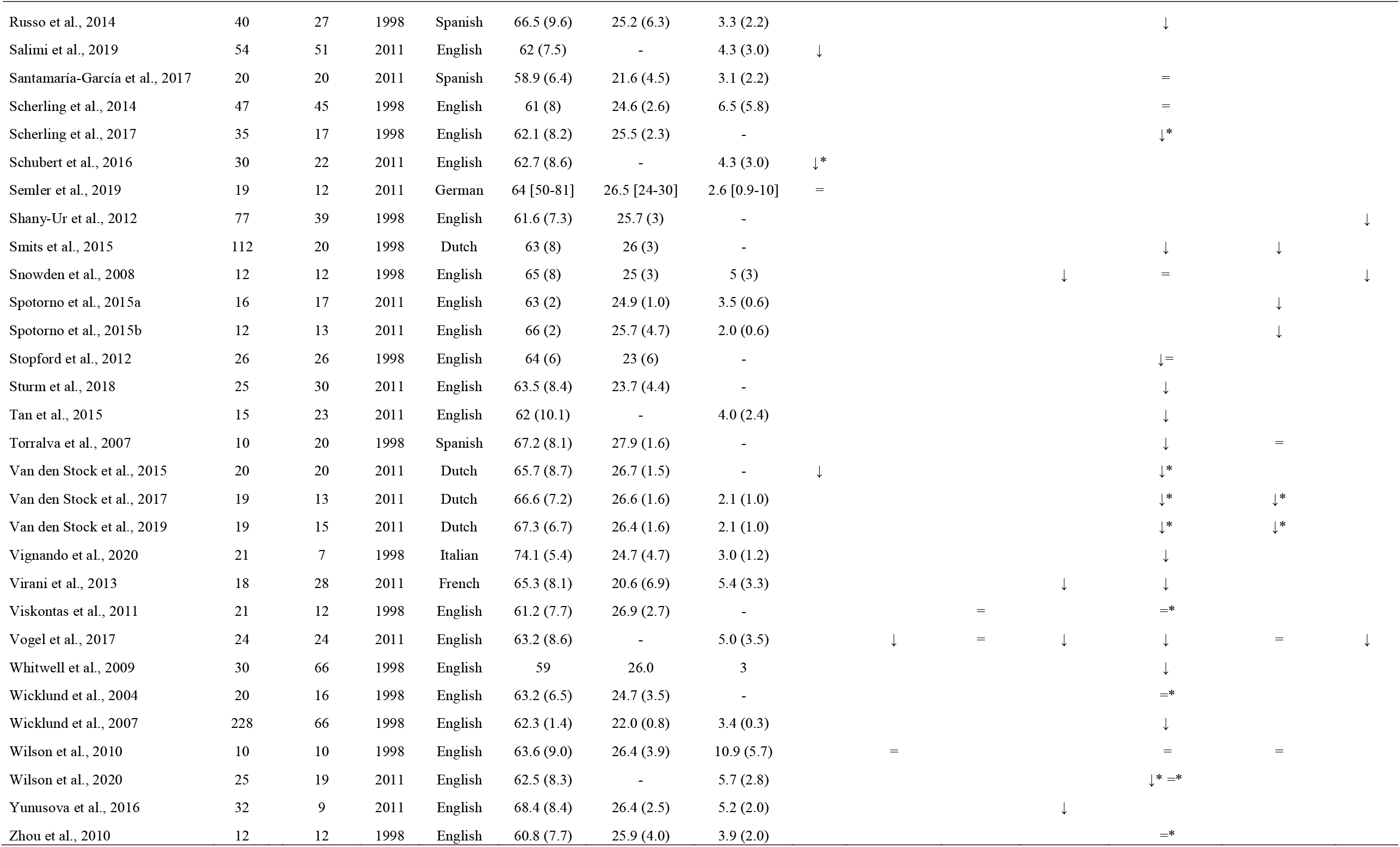
Summary of all included studies comparing bvFTD patients with healthy controls on at least one language measure (Arrow pointing down signifies decreased performance in bvFTD versus healthy controls; Equal sign signifies no significant difference versus healthy controls; Both arrow pointing down and equal sign signifies mixed results; Asterisk signifies that article was not described in the text since another study from the same group and same language measure had a larger sample size).

**Figure 1:**
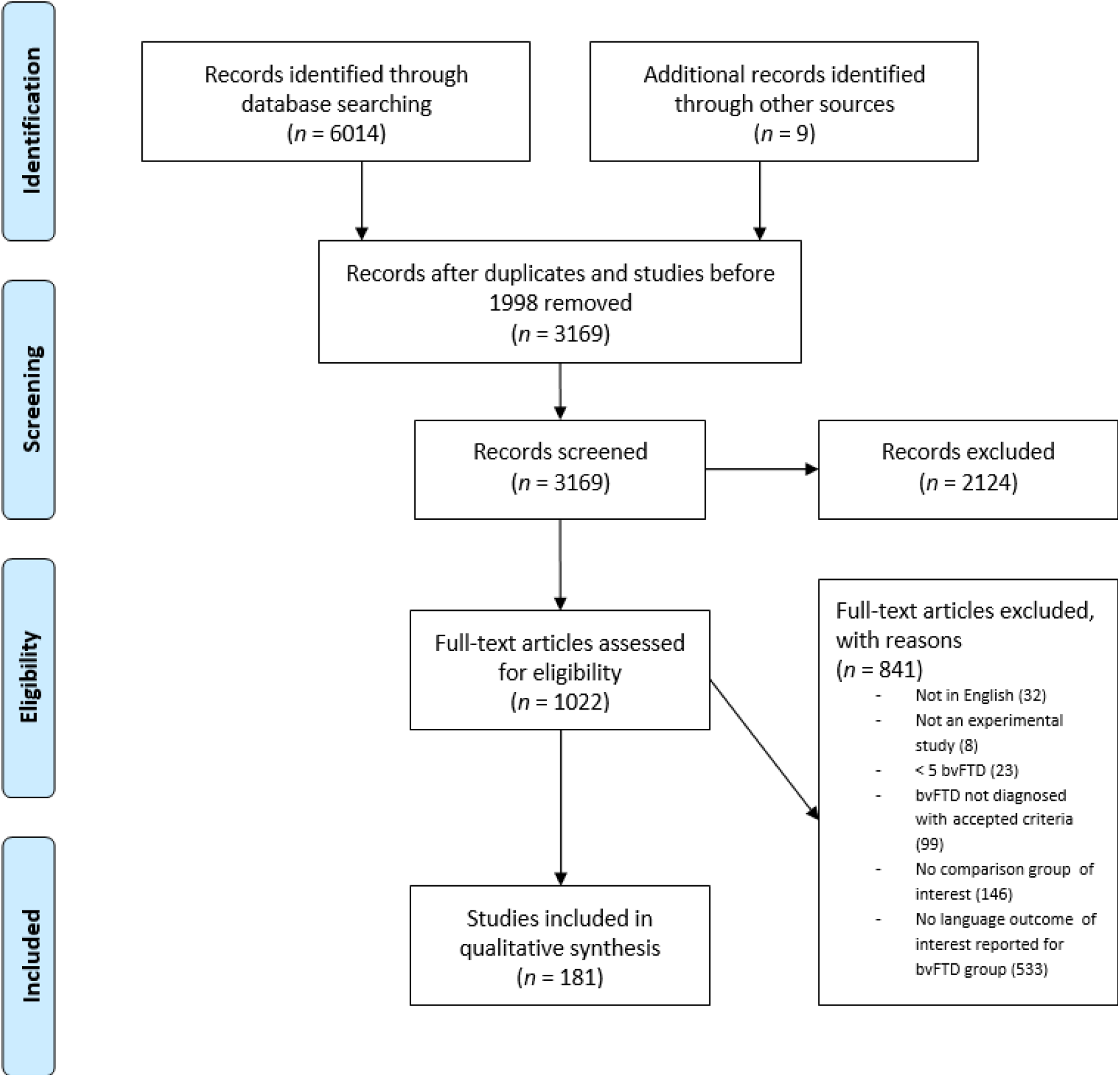
PRISMA Flowchart of literature search.

**Figure 2:**
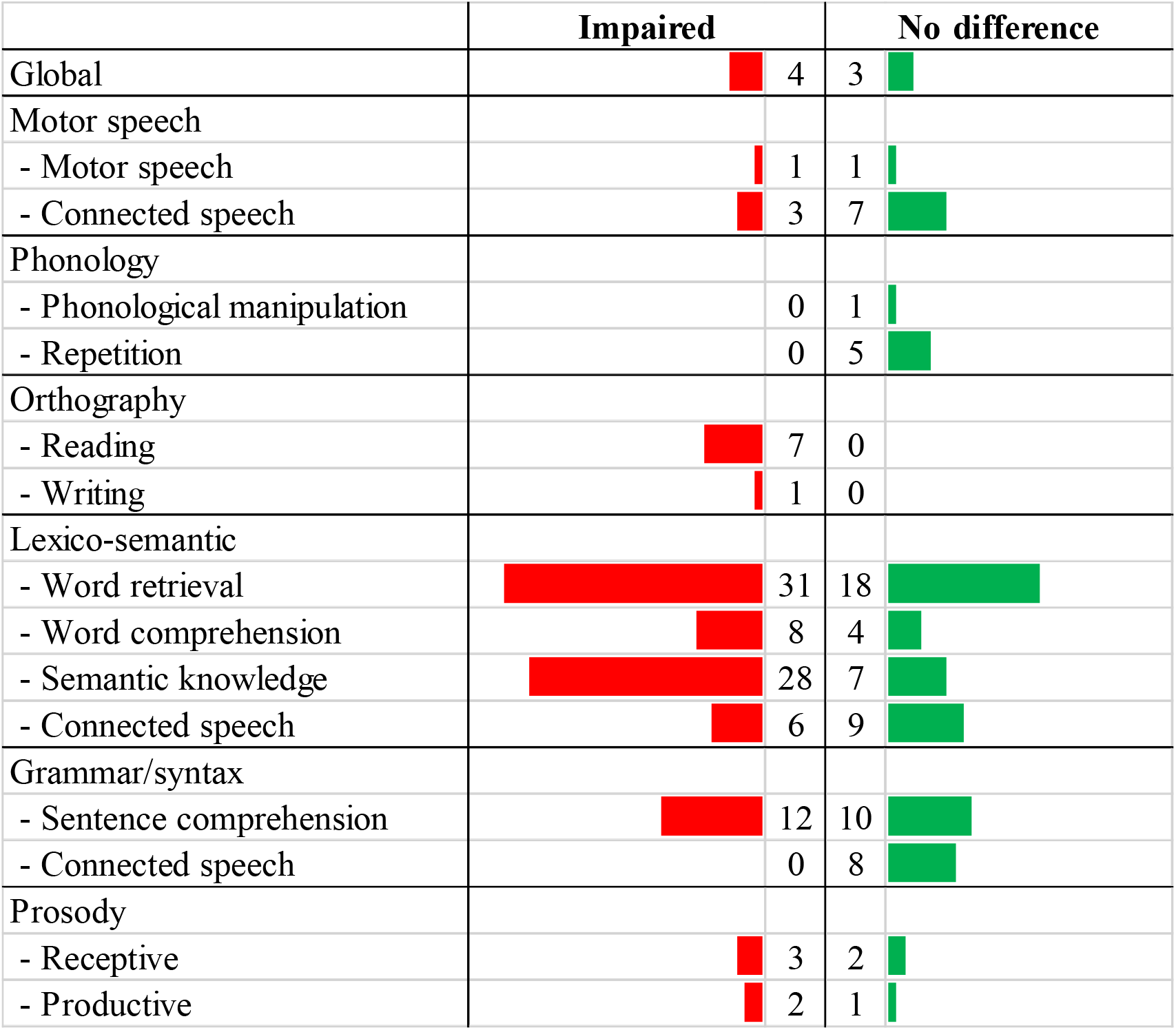
Summary of impaired and preserved language domains in bvFTD, in comparison to healthy controls. For each language subdomains, the number of studies in which the subdomain was altered (red bars) or spared (green bars) is indicated.

### 2.4. Study quality assessment

The risk of bias in studies was assessed at the study level with an adapted version of the Newcastle Ottawa Scale (Wells et al., 2000), recommended for non-randomized studies, including case-control and cohort studies. We assessed each study for (i) sample representativeness, (ii) sample size, (iii) comparability between patients and controls, (iv) ascertainment of language symptoms, and (v) statistical quality (Supplementary material). For the interpretation of the total scores, ranging from 0 to 5, studies were judged to be of low risk of bias (≥ 3 points) or high risk of bias (< 3 points). Therefore, a higher score on this tool represents a lower risk of bias.

## 3. Results

### 3.1. Impaired and preserved language domains in bvFTD

From the 181 selected studies (Figure 1), 156 compared bvFTD to healthy control participants on at least one language measure (Table 1, Figure 2).

#### 3.1.1. Global language performance

Language has been globally evaluated through measures from diverse batteries: the language subscale of the Addenbrooke Cognitive Examination Revised (ACE-R; including naming, comprehension, repetition, reading, and writing), the Aachen Aphasia Test (including detailed evaluation of spontaneous language, token tests, repetition, writing, naming and comprehension), the Language Aphasia Screening Test (LAST) (including picture naming, repetition, counting from 1 to 10, word-to-picture matching, and comprehension of instructions with three levels of complexity), the Sydney Language Battery (including naming, word-picture matching, picture-to-picture matching, and word repetition tasks), and the Consortium to Establish a Registry for Alzheimer’s Disease (CERAD) language subtest (including verbal fluency and picture naming).

A global language impairment was found in bvFTD when using some global language batteries (language subscale of ACE-R: Lima-Silva et al., 2015; Oliver et al., 2015; Salimi et al., 2019, Aachen aphasia test: Van den Stock et al., 2015). However, other batteries failed to find any difference in bvFTD compared to controls (LAST: Johnen et al., 2018, the Sydney Language Battery: Hsieh et al., 2013 and the language subsection of CERAD: Semler et al., 2019). These mixed results may reflect differences in sample sizes between studies (Hsieh et al., 2013; Semler et al., 2019) or difference in the tasks included in these batteries (for example, LAST do not include reading and writing assessment but ACE-R language do).

#### 3.1.2. Motor speech

Motor speech encompasses diverse mechanisms involved in planning, programming, controlling, coordinating, and executing speech production. Various aspects of this domain (e.g., voice sound, pronunciation, articulation, speech errors) can be assessed through highly controlled tasks or through connected speech.

##### 3.1.2.1. Motor speech examination

Motor speech as assessed using controlled tasks seems relatively preserved in bvFTD. Vogel et al. extensively characterized motor speech signatures in 24 bvFTD using perceptual and acoustical analyses of tasks from the Progressive Aphasia Language Scale (saying the day of the week, sequential motion rate and alternating motion rate tasks (i.e., repeating /papa/ or /pataka/), pronouncing a sustained vowel). Objective and listener-based analysis showed that while motor speech is globally preserved, two-thirds of bvFTD showed more subtle motor speech changes (i.e., shorter phrases, longer pauses, strangled-strained voice and articulation difficulties, impaired diadochokinetic performance) (Vogel et al., 2017). Mandelli et al. also showed preserved verbal agility (Mandelli et al., 2016).

##### 3.1.2.2. Motor speech through connected speech

Despite partly contradictory evidence (Vogel et al., 2017), speech rate seems globally preserved in bvFTD (Ash et al., 2013; Gunawardena et al., 2010; Pressman et al., 2019; Wilson et al., 2010). Indeed, even in the infrequent reports showing decreased speech rate, this deficit disappears upon excluding long pauses (Ash et al., 2016). Consistently, compared with controls, bvFTD patients produce more frequent, variable, and extended silent pauses (Nevler et al., 2017; Vogel et al., 2017; Wilson et al., 2010), with no differences in the number of filled pauses and shorter speech segments (Nevler et al., 2017; Vogel et al., 2017). However, the rate of global speech errors (Ash et al., 2009, 2019; Gunawardena et al., 2010) and phonemic errors (Ash et al., 2013), as well as phonetic errors and distortions (Ash et al., 2013; Wilson et al., 2010) is similar between bvFTD patients and healthy persons.

#### 3.1.3. Phonology

The term phonology refers to abstract categories capturing partial recurrent similarities between speech sounds. Phonology may be evaluated through sound manipulation or word/sentence repetition tasks.

##### 3.1.3.1. Phonological manipulation

bvFTD patients showed preserved phonological abilities when assessed with minimal pairs discriminations of the PALPA3 (i.e. discriminating pairs of words that differ in a single phonemic consonant, for example, ‘cut’ and ‘gut’) (Hardy et al., 2016).

##### 3.1.3.2. Repetition tasks

Repetition capacities are preserved for both words and sentences in bvFTD (Chen et al., 2018; Hardy et al., 2016; Mandelli et al., 2016; Viskontas et al., 2011; Vogel et al., 2017).

#### 3.1.4. Orthography

Orthography refers to a system of conventions for reading or writing a language.

Reading appears impaired in bvFTD. Most studies used the National Adult Reading Test (NART), assessing irregular single word reading (Downey et al., 2015; Gregory et al., 2002; Snowden et al., 2008; Virani et al., 2013), and one study focused on non-word reading (Hardy et al., 2016). Decreased text-reading abilities have also been shown in bvFTD patients, with slower articulation rate, elevated number of pauses, and finally, longer and more variable pauses (Vogel et al., 2017; Yunusova et al., 2016).

Writing, as investigated with the Graded difficulty Spelling Test (assessing spelling of orthographically ambiguous words which could plausibly be written in more than one way while retaining the same sound) is also impaired in bvFTD patients (Hardy et al., 2016).

#### 3.1.5. Lexico-semantics

Lexico-semantics refers to the mapping of canonical conceptual information onto particular lexical units. This can be measured in both production and comprehension, with dedicated tasks tapping on sub-domains such as word retrieval, word comprehension, and semantic knowledge. Lexico-semantics can also be assessed more ecologically through connected speech tasks.

##### 3.1.5.1. Word retrieval

Naming, as assessed using traditional tasks, is most frequently impaired in bvFTD patients. The most frequently used task was the Boston Naming Test (BNT), with 12 studies revealing impairments (Ash et al., 2016; Banks & Weintraub, 2008; Couto et al., 2013; De Winter et al., 2016b; Fernandez-Duque & Black, 2007; Lin et al., 2016; Peelle et al., 2008; Russo et al., 2014; Tan et al., 2015; Torralva et al., 2007; Whitwell et al., 2009; Wicklund et al., 2007) and six reporting preserved abilities (Chan et al., 2015; Gleichgerrcht et al., 2012; Kamminga et al., 2014; McKinnon et al., 2008; Rankin et al., 2007; Scherling et al., 2014). Overall, longer versions of the BNT (i.e., 30 or 60 items) seem more powerful to reveal deficits than shorter versions (i.e., 15 or 20 items), which provide mixed results. Using the DO-80 task, two studies showed impairment with the full-length version (Matuszewski et al., 2006; Piolino et al., 2007) but one study using the abbreviated version did not show any differences between the two groups (Rousseaux et al., 2010). Studies using the Graded Naming Test (GNT) provided mixed results, ranging from impaired (Downey et al., 2015; Gregory et al., 2002) to preserved (Snowden et al., 2008) performance.

Additional evidence suggests that naming deficits in bvFTD may be driven by specific lexical categories. Hardy et al. investigated verb naming and revealed a decreased performance bvFTD (Hardy et al., 2016). Indeed, action naming may be impaired despite preserved object naming abilities in bvFTD (Cotelli et al., 2006). As for famous face naming (Cotelli et al., 2006), results are mixed as some studies showed altered capacities (Clague et al., 2005; Kamminga et al., 2015) whereas others found no differences (Clague et al., 2005; Keane et al., 2002). Finally, studies using non-verbal stimuli also provided mixed findings: two studies of odor naming provided diverging results (Luzzi et al., 2007; Orasji et al., 2016), while studies of sound naming showed no difference in comparison with healthy controls (Chow et al., 2010; Lin et al., 2016).

##### 3.1.5.2. Word comprehension

Word comprehension, as assessed using word-picture matching tasks, appears impaired in bvFTD (Chen et al., 2018; Cotelli et al., 2006; Downey et al., 2015; Rhee et al., 2001; Rogers et al., 2006; Sturm et al., 2018; Vignando et al., 2020; Vogel et al., 2017), although other studies did not show any difference between bvFTD and healthy controls (Irish et al., 2011; Luzzi et al., 2007; Miller et al., 2012; Stopford et al., 2012). Of note, these studies included fewer patients, and with shorter disease duration (Table 1). Patients with bvFTD were also impaired on a word definition task (Downey et al., 2015).

##### 3.1.5.3. Verbal semantic knowledge

Verbal semantic processing through semantic associations (matching two semantically-linked words together) is most frequently impaired in bvFTD (Cousins et al., 2016; Rogers et al., 2006 vs. Cosentino et al., 2006 with only 12 patients in this study).

Semantic knowledge (as assessed through the categorical or attribute knowledge of objects or public events) is also decreased in bvFTD (Irish et al., 2016; Johnson et al., 2011; Matuszewski et al., 2006). Deficits have also been revealed through tasks tapping on analogy processing (Krawczyk et al., 2008; Morrison et al., 2004) as well as proverb or idiom comprehension (Luzzi et al., 2020; Reyes et al., 2018). Semantic categorization (defining which item belongs to a target category) is also impaired in bvFTD (Hughes et al., 2011; Koenig et al., 2006 vs. Grossman et al., 2004). As for synonyms matching task, results are more mixed: some studies show impaired performance (Fletcher et al., 2016) while others show preserved abilities (Hsieh et al., 2012). More specifically, Hardy et al. showed that performance is spared for abstract synonyms and impaired for concrete synonyms in these patients (Hardy et al., 2016). Emotional semantics is also impaired in bvFTD (Eckart et al., 2012; Hsieh et al., 2012). Finally, semantic difficulties in bvFTD also impact more global tasks like judging or understanding the organization of actions in scripts (for example, for ‘going fishing’: ‘open can of worms / place worm on hook’) (Cosentino et al., 2006; Farag et al., 2010).

##### 3.1.5.4. Non-verbal semantic knowledge

Although not a linguistic domain per se, non-verbal semantic knowledge is a critical domain to assess in parallel with verbal semantic knowledge. This allows disentangling the extent to which deficits in lexico-semantic tasks are driven by lexical (e.g. word category) vs. conceptual (e.g. semantic features) factors. Non-verbal semantic knowledge is most frequently impaired in bvFTD patients when assessed using pictorial semantic associations (matching two semantically associated pictures) (Ash et al., 2013; Gleichgerrcht et al., 2012; Gunawardena et al., 2010; Rogers et al., 2006; Tan et al., 2015; Torralva et al., 2007 vs. McKinnon et al., 2008; Orasji et al., 2016 with less than nine patients in each of these studies). Semantic knowledge of odours is also globally impaired in bvFTD (Luzzi et al., 2007; Magerova et al., 2014; Orasji et al., 2016 vs. Omar et al., 2013) without difference for flavors (Omar et al., 2013). Studies on semantic knowledge for sounds provide mixed results: some showed impairments (Clark et al., 2017; Fletcher et al., 2015, 2016) while others revealed no difference compared to controls (Chow et al., 2010; Lin et al., 2016). Semantic knowledge for people (face-name matching tasks) are also mixed: one study revealed lower scores in bvFTD (De Winter et al., 2016b) while two studies failed to reveal any difference with controls (Clague et al., 2005; Keane et al., 2002).

##### 3.1.5.5. Lexico-semantics through connected speech

Lexico-semantic abilities using connected speech provide mixed results in bvFTD. Word-level measures are comparable to healthy controls, while discourse-level measures are typically impaired.

In terms of word-level features, the use of lexical categories (nouns, pronouns, verbs, open-class words, closed-class words) does not differ between bvFTD and controls (Ash et al., 2009, 2013, 2016; Hardy et al., 2016; Wilson et al., 2010), except for quantifiers, which occur less commonly in bvFTD patients’ discourse (Ash et al., 2016). Average lexical frequency of the words and nouns used is also comparable to healthy controls (Hardy et al., 2016; Wilson et al., 2010). Considering errors, results are mixed for semantic paraphasias and word-finding difficulties (Ash et al., 2006; Wilson et al., 2010: no difference vs. Rousseaux et al., 2010: more frequent).

In terms of discourse-level features, accuracy of the discourse (as judged through global organization, complexity or the ability to remain on topic) seems to be preserved when investigated with a global index (Gola et al., 2015; Mendez et al., 2014), but more fine-grained examinations reveal impairments in accurately reporting events and precisely guiding communication (with insufficient or superfluous responses) (Ash et al., 2006; Healey et al., 2019; Healey et al., 2015). As for connectedness, even if measures of action or evaluation clauses and temporal organization revealed no difference (Gola et al., 2015), other studies revealed impairments in global connectedness, maintenance of a theme and discourse organization (Ash et al., 2006; Rousseaux et al., 2010).

#### 3.1.6 Grammar

Grammar refers to the system of Sequencing of hierarchically organized morphosyntactic patterns, determining the well-formedness or ill-formedness of sentences and words. It underlies both sentence comprehension and production (although no study investigated sentence production in bvFTD). Grammatical features can also be extracted from connected speech.

##### 3.1.6.1. Sentence comprehension

Overall, in bvFTD, sentence comprehension was most frequently preserved when using simple sentence comprehension tasks, but impaired when using more complex experimental sentence comprehension assessments.

Most studies on simple sentence comprehension employed sentence-picture matching tasks (selecting which of many pictures matches a given sentence). Four of them showed preserved abilities (Cotelli et al., 2007; Hardy et al., 2016; Johnson et al., 2011; Rosen et al., 2004) and two showed decreased scores (Charles et al., 2014; Mandelli et al., 2016). Other studies used instructions tasks (e.g., asking the patient to follow instructions that are variable in length and linguistic complexity such as ‘Point to a square’ or ‘Put the small red square on the large blue circle’). Three showed preserved performance (Gleichgerrcht et al., 2012; Torralva et al., 2007; Vogel et al., 2017) and one showed impaired abilities (Giovagnoli et al., 2008). Finally, some studies assessed sentence comprehension using comprehension questions on sentences: two showed preserved abilities (Ash et al., 2009; Peelle et al., 2008) and one revealed difficulties with comparative questions (Smits et al., 2015).

Contrary to simple sentence comprehension tasks, those employing more complex materials have mainly revealed deficits, such as difficulties during a discourse task (Grossman et al., 2005; Luzzi et al., 2020; Rousseaux et al., 2010) or with ambiguous sentences (McMillan et al., 2013; Spotorno et al., 2015a). Other authors have found a partial sensitivity to syntactic violations (Murray et al., 2007; Peelle et al., 2007). For example, patients were impaired in grammatically judging an unknown verb used with correct or violated grammatical rules (Murray et al., 2007). They were also less sensitive to syntactic errors (as, for example, ‘The book is being closely pick/picked by the group of curious students’ or ‘The friend is being reunion/met in the shopping mall entrance’) (Peelle et al., 2007).

##### 3.1.6.2. Grammar through connected speech

Connected speech analyses reveal preserved grammar in bvFTD patients (Ash et al., 2013, 2016, 2019; Gunawardena et al., 2010; Rousseaux et al., 2010; Vogel et al., 2017; Wilson et al., 2010). More precisely, patients do not significantly differ from controls in utterance length (Ash et al., 2013; Wilson et al., 2010), word and clause complexity (Ash et al., 2013, 2016, 2019; Gunawardena et al., 2010; Wilson et al., 2010), and correct and incorrect sentences (Ash et al., 2013; Rousseaux et al., 2010; Vogel et al., 2017; Wilson et al., 2010).

#### 3.1.7. Prosody

Prosody is a supra-segmental feature of language using modulation of intonation, rhythm, and intensity to convey emotional or linguistic distinctions.

Receptive prosody appears impaired in bvFTD. The most frequently used tasks are emotion labelling tasks, in which the participant must listen to stimuli with distinct prosodic features and pick an emotional label (e.g., anger, sadness, happiness). Most studies revealed global impaired recognition of prosody using vocal signals or sentences (Keane et al., 2002; Shany-Ur et al., 2012; Snowden et al., 2008), although two studies did not find any significant difference in bvFTD patients (Omar et al., 2011; Rankin et al., 2009). Another study suggested that this impairment in bvFTD may depend on emotion type: impaired recognition of happiness, sadness, surprise, and anger but preserved recognition of fear and disgust (Keane et al., 2002).

Fewer studies have assessed prosody during speech production in bvFTD. These studies showed reduced stress (Vogel et al., 2017), mixed results for pitch modulation (altered in Nevler et al., 2017 but preserved in Pressman et al., 2019 and Vogel et al., 2017), and preserved intensity (Pressman et al., 2019).

### 3.2. Language profiles according to bvFTD subgroups

From the 181 studies assessing language in bvFTD (Figure 1), 26 compared patient subgroups divided upon four criteria (Table 2): clinical, neural, genetic or neuropathological subgroups. Such comparisons aim to identify which features may be more associated with language impairments and potentially informing on the heterogeneity of language profiles across the bvFTD population.

**Table 2.** Summary of all included studies comparing bvFTD subgroups on at least one language measure.

| Author, year | bvFTD subgroups (n) | Language results | Criteria | Language |
| --- | --- | --- | --- | --- |
| Bertoux et al., 2014 | -Non-amnesic bvFTD (20)<br>-Amnesic bvFTD (24) | Picture naming did not differ between amnesic and non-amnesic bvFTD. | 2011 | French |
| Bertoux et al., 2016 | -Non-amnesic bvFTD (19)<br>-Amnesic bvFTD (19) | Picture naming did not differ between amnesic and non-amnesic bvFTD. | 2011 | French |
| Cerami et al., 2016 | -bvFTD frontal (25)<br>-bvFTD temporo-limbic (27) | Instruction comprehension was more impaired in bvFTD frontal than in temporo-limbic predominant. | 2011 | Italian |
| Deutsch et al., 2015 | -bvFTD without traumatic brain injury (581),<br>-bvFTD with traumatic brain injury (25) | Picture naming did not differ between bvFTD with or without traumatic brain injury | 1998 | English |
| Downey et al., 2014 | -bvFTD-sporadic (5)<br>-bvFTD-C9 (5)<br>-bvFTD-MAPT (7) | Single word comprehension did not differ between the three bvFTD subgroups. Picture naming scores were higher in bvFTD-C9 than in bvFTD-MAPT and controls. | 2011 | English |
| Fernández-Matarrubia et al., 2017 | -Non-amnesic bvFTD (13)<br>-Amnesic bvFTD (13)<br>- CTRL (24) | Picture naming did not differ between amnesic and non-amnesic bvFTD. | 2011 | Spanish |
| Gleichgerrcht et al., 2011 | -Non-utilitarian bvFTD (13)<br>-Utilitarian bvFTD (9) | No difference in picture naming or comprehension of instructions between utilitarian and non-utilitarian bvFTD profiles. | 1998 | Spanish |
| Hardy et al., 2016 | -bvFTD-MAPT (6)<br>-bvFTD-C9 (4)<br>-CTRL (24) | Word comprehension and picture naming were lower in bvFTD-MAPT than bvFTD-C9. Concrete and abstract synonyms as well as writing scores were lower in bvFTD-MAPT than in bvFTD-C9. Verb naming, words and non-words reading, auditory processing, words repetition and sentence-picture matching did not differ between bvFD-MAPT and bvFTD-C9. | 2011 | English |
| Heuer et al., 2020 | -bvFTD (135)<br>-bvFTD familial (99)<br>-bvFTD-C9 (43)<br>-bvFTD-GRN (14)<br>-bvFTD-MAPT (28) | Regular words, irregular words, sentence reading and sentence repetition did not differ between the bvFTD subgroups. | 2011 | English |
| Kipps et al., 2009 | -bvFTD MRI abnormal (15)<br>-bvFTD MRI normal (9)<br>-CTRL (10) | Picture naming did not differ between bvFTD with normal or abnormal MRI. | 1998 | English |
| Kramer et al., 2018 | -bvFTD with abnormal PET (20)<br>-bvFTD with normal PET(10) | Naming did not differ between bvFTD with a normal or abnormal FDG-PET | 2011 | German |
| Manes et al., 2011 | -bvFTD with impaired neuropsychologic performance (30)<br>-bvFTD with normal neuropsychologic performance (13)<br>-CTRL(14) | Picture naming scores were lower in bvFTD with impaired neuropsychologic performance than in ones with normal neuropsychologic performance. | 1998 | Spanish |
| Massimo et al., 2019 | -bvFTD professionals (50)<br>-bvFTD non professionals (13) | Picture naming performance did not differ between professionals and non professionals bvFTD. | 2011 | English |
| Mendez et al., 2017 | -bvFTD PvSp + (9)<br>-bvFTD PvSp – (3) | Pressure of speech, tangeantiality, derailment and clanging were more frequent in bvFTD with poverty of speech than without, but the amount of neologisms, word approximations and perseverations, echolalia or blocking, the poverty of the content of speech, its incoherence, illogicality and loss of goal did not differ between the two bvFTD subgroups | 2011 | English |
| Mendez & Lim, 2004 | -right bvFTD (8)<br>-left bvFTD (8)<br>-CTRL (24) | Picture naming was lower in right bvFTD compared to left bvFTD | 1998 | English |
| Mendez & Shapira, 2013 | -bvFTD-FTLD (74)<br>-bvFTD-AD (21)<br>-bvFTD-tau negative (51) | Picture naming did not differ between bvFTD-FTLD and bvFTD-AD, nor between bvFTD-tau positive and bvFTD-tau negative | 1998;<br>2011 | English |
|  | -bvFTD-tau positive (23) |  |  |  |
| Moore et al., 2020 | -bvFTD-MAPT (15)<br>-bvFTD-C9 (56)<br>-bvFTD-GRN (33)<br>-CTRL (248) | Semantic association scores did not differ between bvFTD due to a MAPT mutation and bvFTD-C9 or bvFTD-GRN. | 2011 | English |
| Park et al., 2017 | -bvFTD with parkinsonism (33)<br>-bvFTD without parkinsonism (38) | A global language score did not differ between bvFTD with or without parkinsonism. | 2011 | Korean |
| Perry et al., 2017 | - bvFTD-AD (7)<br>- bvFTD-FTLD (50)<br>- bvFTD-FTLD-tau (15)<br>- bvFTD-FTLD-TDP (31)<br>-bvFTD-FTLD-FUS (4)<br>- bvFTD-CBD (6)<br>- bvFTD-PSP (5)<br>- bvFTD-TDP-A (5)<br>- bvFTD-TDP-B (15)<br>- bvFTD-TDP-C (5)<br>- bvFTD-TDP-U (6)<br>- bvFD-aFTLDu (4) | Repetition scores were lower in bvFTD-AD than in bvFTD-FTLD and did not differ between bvFTD-FTLD subgroups. Semantic association scores were lower in bvFTD-TDP B than in bvFTD-TDP U. Picture naming scores were lower in bvFTD-TDP C than in bvFTD linked to CBD, TDP U and aFTLD-U. Verbal agility did not differ between the bvFTD subgroups. | 2011 | English |
| Ranasinghe et al., 2016 | -bvFTD Salience Network Frontal-Temporal (SN-FT) (21)<br>-bvFTD Semantic Appraisal Network (SAN) (8)<br>-bvFTD Salience Network Frontal (SN-F) (27)<br>-bvFTD subcortical (30) | Syntax comprehension scores were lower in bvFTD SN-FT than in bvFTD SN-F and bvFTD subcortical. Picture naming scores were lower in bvFTD SAN than in bvFTD SN-F and bvFTD subcortical. Verbal agility, affect naming, reading irregular words and repetition did not differ between the bvFTD subgroups. | 2011 | English |
| Roca et al., 2013 | -Low-functioning bvFTD (19)<br>-High-functioning bvFTD (16)<br>-CTRL (14) | Picture naming scores were lower in low-functioning bvFTD when compared to high-functioning bvFTD and controls. Semantic associations were impaired in low-functioning bvFTD when compared to controls but the other subgroups. | 1998 | Spanish |
| Sha et al., 2012 | -bvFTD without C9 (48)<br>-bvFTD-C9 (15) | Syntax, naming, reading and repetition did not differ between bvFTD-C9 and bvFTD without C9 | 2011 | English |
| Suhonen et al., 2017 | -bvFTD without C9 (44)<br>-bvFTD-C9 (16) | Picture naming and semantic processing did not differ between bvFTD-C9 and bvFTD without C9 expansion. | 2011 | Finnish |
| Torralva et al., 2009 | -bvFTD with low score on Addenbrooke Cognitive Examination (19)<br>-bvFTD with high score on Addenbrooke Cognitive Examination (16)<br>-CTRL (14) | Picture naming was lower in bvFTD with low score on Addenbrooke Cognitive Examination than in bvFTD with high score on Addenbrooke Cognitive Examination. Instructions comprehension was lower bvFTD with low score on Addenbrooke Cognitive Examination than in bvFTD with high score on Addrenbroke Cognitive Assessment. | 1998 | Spanish |
| Torralva et al., 2015 | -Non-vascular bvFTD (66)<br>-Vascular bvFTD (23) | Picture naming did not differ between vascular and non-vascular bvFTD. | 1998 | Spanish |
| Ye et al., 2015 | -bvFTD early onset (47)<br>-bvFTD late onset (29) | Picture naming did not differ between early and late onset bvFTD. | 2011 | Korean |

#### 3.2.1. Comparisons between clinically defined bvFTD subgroups

Few clinical factors influencing language profiles in bvFTD have been identified so far. First, bvFTD patients with impaired global cognitive performance had more severe deficits in picture naming (Manes et al., 2011; Roca et al., 2013), sentence comprehension (Torralva et al., 2009) and semantic knowledge (Roca et al., 2013). Furthermore, patients with poverty of speech had more pressure of speech, tangentiality (i.e. digressions, free associations of ideas), derailment, and clanging (associations of words based upon sounds) than those without this feature (Mendez et al., 2017). The level of confidence of the diagnosis (probable vs. possible bvFTD) provided conflicting results. While Kipps et al. found a difference in picture naming in patients with a diagnosis of probable bvFTD versus possible bvFTD (Kipps et al., 2009), Borroni et al. and Kramer et al. did not find any significant difference in the language profile of probable and possible bvFTD patients (Borroni et al., 2015; Kramer et al., 2018).

None of the other investigated clinical factors had an impact on language measures. Global language skills do not seem affected by the presence of absence of parkinsonism (Park et al., 2017). Naming abilities are not influenced by the presence or absence of memory deficits (Bertoux et al., 2014, 2016; Fernández-Matarrubia et al., 2017), moral judgment profiles (Gleichgerrcht et al., 2011) or a medical history of vascular disease (Torralva et al., 2015) or traumatic brain injury (Deutsch et al., 2015). Finally, sentence comprehension seems unaffected by age at onset (Ye et al., 2015) or profession (Massimo et al., 2019).

#### 3.2.2. Comparisons between neuroanatomically defined bvFTD subgroups

First, a frontal dominant vs. temporo-limbic dominant atrophy pattern seems to impact sentence comprehension. Frontal predominant bvFTD show more difficulties in the Token task than temporo-limbic predominant bvFTD (Cerami et al., 2016).

The laterality of atrophy, or right vs. left predominant atrophy, significantly affects naming abilities, with left-lateralized bvFTD patients showing lower performance than right-lateralized cases (Mendez & Lim, 2004).

Finally, one study investigated differences between bvFTD subgroups based on their patterns of network degeneration (with a predominance in the semantic appraisal network (SAN), in the salience network (SN), subdivided in frontal (SN-F) and fronto-temporal (SN-FT) predominant subgroups, or in the subcortical network (subcortical)). While affect naming did not differ between all the subgroups, picture naming was lower in the SAN bvFTD subgroup. The SN-FT bvFTD subgroup had the second lowest scores without significant difference with other groups. Sentence comprehension was lower in the SN-FT subgroup, differing significantly from SN-F and subcortical subgroups. The SAN bvFTD subgroup had the second lowest scores, without significant difference with other groups (Ranasinghe, 2016). Irregular words reading, repetition as well as verbal agility did not differ between all the subgroups (Ranasinghe et al., 2016).

#### 3.2.3. Comparisons between genetically defined bvFTD subgroups

First, a global genetic origin (familial bvFTD vs. sporadic bvFTD) does not affect repetition and reading abilities (Heuer et al., 2020).

More specifically, relative to patients with *C9orf72* expansions, those with *MAPT* mutations exhibit greater deficits in writing, object naming and in two different tasks of single word comprehension (Downey et al., 2014; Hardy et al., 2016). However, other studies reported that the *MAPT* mutations, in comparison to other genetic mutations (*C9orf72* or *GRN*), does not influence performance in repetition, reading (Hardy et al., 2016; Heuer et al., 2020), single-word comprehension (Downey et al., 2014), semantic knowledge (Moore et al., 2020) or sentence comprehension (Hardy et al., 2016) disproportionately.

However, the presence of a *C9orf72* or *GRN* expansion does not lead to more severe language impairments for any of the measures investigated, in comparison to other genetic mutations (Downey et al., 2014; Hardy et al., 2016; Heuer et al., 2020; Moore et al., 2020; Sha et al., 2012; Suhonen et al., 2017).

#### 3.2.4. Comparisons between neuropathologically defined bvFTD subgroups

When comparing bvFTD linked to FTLD vs. AD pathologies, no difference was found in naming (Mendez & Shapira, 2013; Perry et al., 2017) or word comprehension, but repetition scores were lower in AD than in FTLD patients (Perry et al., 2017).

Considering major FTLD molecular class, no language difference was found between FTLD-tau, TDP and FUS (Perry et al., 2017) nor between tau-positive and tau-negative bvFTD patients (Mendez & Shapira, 2013). However, Perry et al. also compared bvFTD linked to different FTLD subtypes: CBD, PSP, FTLD-TDP (type A, B, C, D or unclassified) and FUS, when sample size permitted. They found that patients with a FTLD-TDP type C pathology performed significantly lower in picture naming in comparison to PSP and FTLD-TDP unclassified, but not CBD, FTLD-TDP type A or FTLD-TDP type B. Also, patients with a FTLD-TPD type B performed significantly lower in word comprehension, in comparison to FTLD-TDP unclassified (Perry et al., 2017).

### 3.3. Study quality assessment

Based on our assessment for the risk of bias 130 studies had a low risk of bias and 51 studies had a high risk of bias. Table S1 indicates, for each study, the risk of bias in each item assessed (e.g., representativeness, sample size, comparability, outcome, and statistics). The highest risk of bias was due to small sample size (i.e., less than 20 patients overall), to inadequate comparability of the healthy controls with the bvFTD (i.e., cases and controls were only partially matched on basic demographic variables), and to partial independent validation and/or description of patient selection criteria (Figure 3). Most studies employed validated neuropsychological tools to test speech and language outcomes, while studies without validated measurement tools mainly included experimental tasks and/or measures derived from spontaneous, narrative or connected speech.

**Figure 3:**
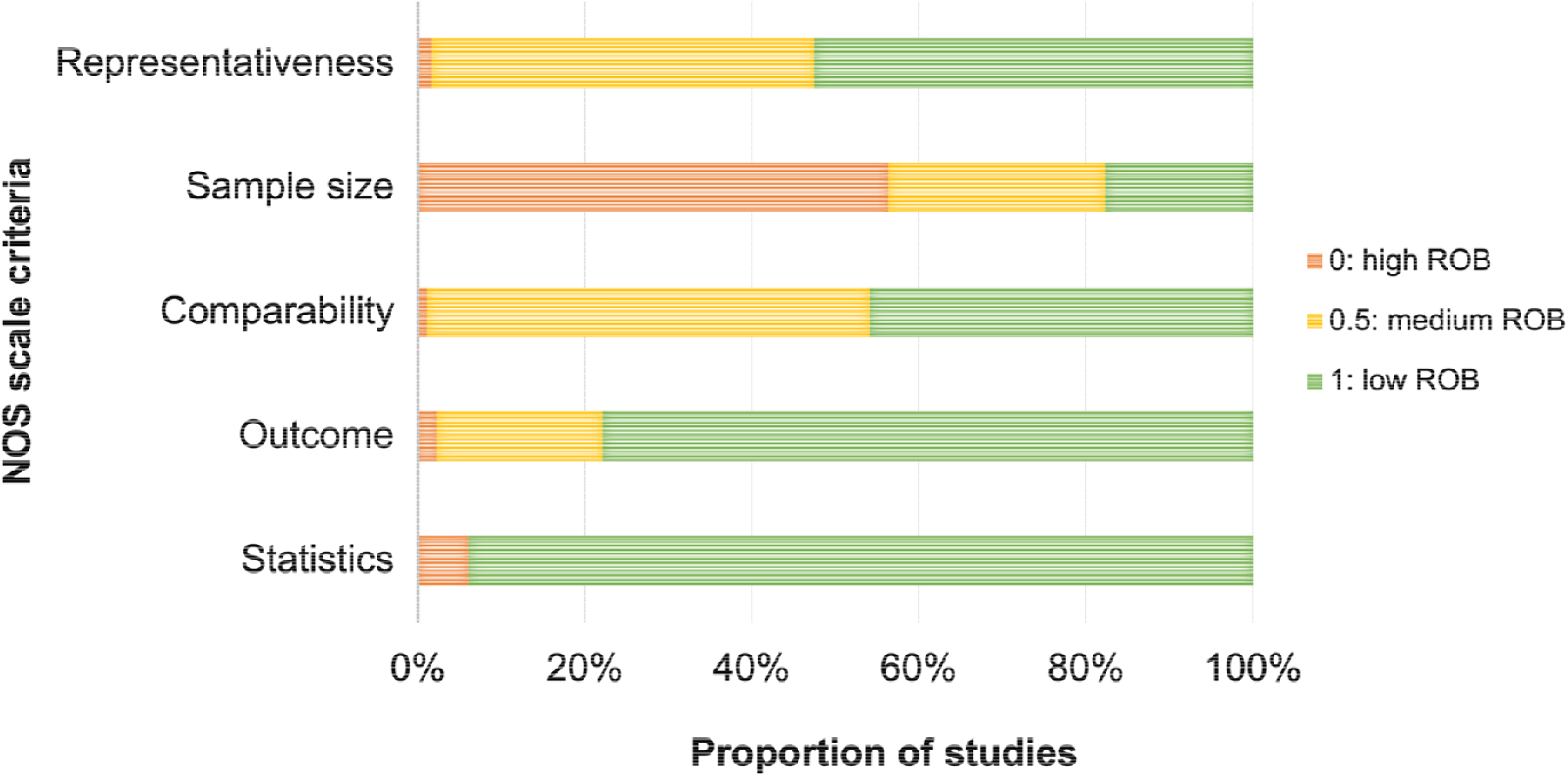
Risk of bias assessment.

## 4. Discussion

In this review, we investigated the scope of speech and language impairments in bvFTD, considering multiple domains (motor speech, phonology, orthography, lexico-semantics, grammar, and prosody) and modulating (clinical, neural, genetic, neuropathological) factors. Overall, we found that bvFTD patients present with extensive but heterogeneous deficits. These were more consistently reported in lexico-semantic, orthographic, and prosodic domains. Assessments of motor speech and grammar provided mixed results, with a slight majority of studies showing preserved performance. Finally, no study showed phonological impairments. Also, speech and language impairments were more pronounced in patients with (i) more severe global cognitive impairment, (ii) probable (as opposed to possible) bvFTD diagnosis, (iii) predominant frontotemporal atrophy (vs. frontal- or subcortical-predominant) or semantic appraisal network damage, and (iv) *MAPT* mutations (vs. *C9orf72*). The impact of underlying neuropathology remains underexplored. Overall, our study suggests that despite being under-recognized, speech and language impairments in bvFTD could afford sensitive biomarkers for both diagnosis and prognosis.

### 4.1. Characterization of language in bvFTD

Three language domains were found to be consistently impaired (more than 60% of reports) in bvFTD: lexico-semantics, orthography, and prosody. First, bvFTD are impaired in the lexico-semantic domains, more precisely in word retrieval, word comprehension and semantic knowledge. Of note, the semantic impairment in bvFTD patients did not appear to be restricted to verbal stimuli but also extended to non-verbal information. However, these lexico-semantic deficits are not always observed in connected speech studies, but fine-grained examinations revealed impairments at the discourse level (connectedness, accuracy, theme maintenance, etc.). In bvFTD patients, lexico-semantic impairments have been related to left anterior temporal lobe and left inferior frontal gyrus (Hardy et al., 2016), which are core regions of the semantic network (Lambon Ralph et al., 2017) and also atrophied in bvFTD patients (Bejanin et al., 2020). Interestingly, lexico-semantic difficulties may be more marked for actions (verbs) than for objects (nouns) in bvFTD (in naming: Cotelli et al., 2006; Hardy et al., 2016; word comprehension: Rhee et al., 2001, semantic knowledge: Bak & Hodges, 2003). This may suggest a larger involvement of frontal lobe and more specifically motor regions (Birba et al., 2017; García et al., 2018), as well as more executive resources in verbs than nouns processing (Bak et al., 2001). Second, reading capacities are also altered in bvFTD patients, which has most frequently been shown for irregular words, but also for non-words and text. Reading irregular words is related to semantic abilities and involves the anterior temporal role (Wilson et al., 2009), a region targeted by neurodegeneration in bvFTD. Nonetheless, more fine-grained studies involving reading different types of words are needed in bvFTD, as well as more studies of writing abilities. Third, prosody recognition was impaired in most reports. Previous studies showed that this function depends on medial and orbitofrontal regions and on anterior temporal poles which are core regions of neurodegeneration (Pichon & Kell, 2013; Schirmer & Kotz, 2006). bvFTD patients also have impaired expressive prosody, which has been linked to left inferior frontal, anterior cingulate, insula, left fusiform and right inferior frontal gyri (Nevler et al., 2017), regions also atrophied in bvFTD and involved in social and behavioral disorders (Massimo et al., 2019). These results highlight the relevance of prosodic markers in bvFTD, especially given the relationship of prosody with socio-emotional behavior, a core impairment in bvFTD (Rascovsky et al., 2011).

Two language domains showed mixed results in bvFTD, with a slight majority of studies showing preserved performance: motor speech and grammar. Motor speech is globally preserved, even if subtle changes appears when more detailly investigated, related to inferior frontal gyrus, insula and right precentral gyrus (Vogel et al., 2017). These regions have been linked to various motor speech aspects in healthy participants: left anterior insula for articulation, SMA and pre-SMA for initiation and execution of speech (Price, 2010). Grammar appears to be globally preserved when assessed with simple comprehension tasks or in its production during connected speech, but more altered when assessed with complex tasks. This might be due to increased executive demands. In a task-based fMRI study of grammar processing, decreased activation in the dorsal inferior frontal cortex has been found in bvFTD patients in comparison to healthy controls (Cooke et al., 2003). Previous studies in healthy controls have also highlighted the role of left frontal regions in grammar (Cooke et al., 2002; Grossman et al., 2002; McMillan et al., 2013).

One language domain showed consistently preserved performance in bvFTD: phonology. This was observed in studies assessing phonology with both phonological manipulation and repetition of words and sentences. These results are in line with the implication of posterior temporo-parietal regions in this language domains, more posterior than the atrophied regions in bvFTD (Forkel et al., 2020).

### 4.2. The heterogeneity of language impairments in bvFTD

We have also identified factors that are associated with more speech and language impairments in bvFTD patients. Unsurprisingly, a more severe global cognitive impairment leads to more alterations in speech and language in bvFTD patients, which highlight the role of speech and language tasks as additional tools that are sensitive to disease progression and severity. The level of certainty of diagnosis (possible versus probable) also affects the severity of language impairments. This may be related to the fact that within individuals with a diagnosis of possible bvFTD, some may not progress to more severe stages of the disease (“bvFTD phenocopy”) (Hornberger et al., 2009). Consistent with our findings, a previous study also found that there was a higher prevalence of stereotypy of speech in bvFTD progressors, versus non-progressors (Hornberger et al., 2009).

The neurodegeneration pattern may also impact language capacities. A left-predominant pattern of atrophy (vs right-predominant) is associated with more severe naming impairments, consistently with the role of the left hemisphere in speech and language (Mendez & Lim, 2004). This pattern is also observed in other neurodegenerative diseases such as left-predominant svPPA patients who show more severe naming impairments than right-predominant svPPA patients (Binney et al., 2016). Frontal (vs. temporo-limbic) structural alterations as well as altered fronto-temporal connectivity were associated with more significant sentence comprehension difficulties. This is consistent with the predominant role of the left frontal regions in syntax processing (Cerami et al., 2016; Schell et al., 2017) even if temporal lobes are also involved in this process (Lukic et al., 2021). A more semantic appraisal network predominant pattern alters naming capacities more importantly (Ranasinghe et al., 2016), which is consistent with the role of bilateral anterior temporal lobes in semantic processing (Lambon Ralph et al., 2017).

Furthermore, our review suggests that the presence of a *MAPT* mutations impact language, more specifically lexico-semantics (naming, single-word comprehension) and writing, more significantly than *C9orf72*. In that specific study, writing was assessed with the Graded Spelling Test, in which all words are orthographically ambiguous, in the sense that they can plausibly be written in multiple ways, while retaining the sound of the target item (Hardy et al., 2016). Therefore, this task also includes a semantic component. The more severe lexico-semantic impairment in *MAPT* vs *C9orf72* bvFTD patients may be related to a greater involvement of anterior temporal lobe, a region highly implicated in lexico-semantics, in bvFTD patients with *MAPT* mutations than with *C9orf72* expansion (Josephs et al., 2007; Mahoney et al., 2012). There was no significant difference when investigating other speech and language domains, nor when including *GRN* mutations in the comparison. Overall, these preliminary results are surprising, given that outside of a diagnosis of bvFTD, *GRN* mutations are the most frequently associated with a speech and language predominant diagnosis of PPA (Saracino et al., 2021). More studies on genetic cases of bvFTD are needed to confirm the differential speech and language profiles.

As for the effect of underlying neuropathology, it remains to be determined due to the low number of studies. Nonetheless, a few differences have been reported. Underlying AD pathology may alter repetition more than FTLD pathology (Perry et al., 2017), in line with the posterior predominance of AD (Braak & Braak, 1991) and the involvement of posterior temporo-parietal regions in repetition (Forkel et al., 2020). FTLD-TDP type C seems to lead to more naming difficulties in comparison to PSP and FTLD-TDP unclassified, but not CBD, FTLD-TDP type A or FTLD-TDP type B. This is consistent with the implication of FTLD-TDP type C in a large majority of svPPA cases and in the left anterior temporal lobe, a core region for naming (Borghesani et al., 2020). Finally, FTLD-TDP type B seems related to more comprehension difficulties in comparison to FTLD-TDP unclassified, consistent with anterior temporal atrophy described in cases of patients with language disturbances related to FTLD-TDP type B (Lee et al., 2020).

In conclusion, the heterogeneity of speech and language symptoms in bvFTD patients is concordant with previous studies assessing the impact of clinical, neuroanatomical, genetic and neuropathological features on such symptoms. In the first aim of this systematic review, we delineated a relatively clear speech and language profile in bvFTD. Nonetheless, the evidence is still too limited to allow clear conclusions for the second aim. Future prospective studies investigating the impact of these features on speech and language symptoms in bvFTD patients will be essential to manage the clinical complexities of this disease at the single-patient level.

### 4.3. Beyond the characterization of patients: Speech and language measures to improve bvFTD phenotyping and to track disease progression in bvFTD

The language characterization of bvFTD is needed for a better understanding of the bvFTD syndrome, but it may also be useful for its differential diagnosis from other neurodegenerative disease, especially PPA and AD. The profile of speech and language imapirments in bvFTD reported in this review, namely an impairment in lexico-semantics and orthography, with preserved phonological and relatively intact motor speech and grammar, resembles the most to the one described in svPPA. Nonetheless, when compared to svPPA, bvFTD patients show better performance in most lexico-semantic tasks, but do not differ in phonology, reading, motor speech and grammar (Ash et al., 2013; Harciarek & Kertesz, 2008; Hardy et al., 2016; Wilson et al., 2010). Furthermore, when compared to lvPPA, bvFTD patients are less impaired in phonology (repetition), naming and syntactic comprehension but have more fluency disruptions in their discourse (Ash et al., 2013; Smits et al., 2015; Wilson et al., 2010). When compared to nfvPPA, bvFTD patients have better performances on tasks involving motor speech, grammar or verb naming, but did not differ in other language domains such as lexico-semantics (Ash et al., 2013; Hardy et al., 2016; Wilson et al., 2010).

Although most studies show that language in bvFTD do not differ from AD (Blair et al., 2007; Diehl et al., 2005; Harciarek & Kertesz, 2008; Smits et al., 2015; Wilson et al., 2010), it was reported that semantic and motor speech abilities are more impaired in bvFTD than in AD (Blair et al., 2007). Finally, other studies have also shown that speech and language can be helpful in the differential diagnosis of bvFTD versus ALS/FTD (Saxon et al., 2017), CBS and PSP (Cotelli et al., 2006), DLB and vascular dementia (Smits et al., 2015). Thus, a detailed language investigation seems to allow distinctions between bvFTD and other neurodegenerative disorders.

Although only a few studies have assessed speech and language longitudinally in bvFTD, it is of great interest to know which domains could track disease progression. Global language performances seem to decrease with time (Blair et al., 2007; Schubert et al., 2016). A connected speech study also suggests that syntactic complexity and speech rate declines with time in bvFTD (Ash et al., 2019). As for naming, results are mixed (no change with time in (Ash et al., 2019; Binney et al., 2017; Cousins et al., 2018 vs. decrease in Hardy et al., 2016; Mahoney et al., 2014; Wicklund et al., 2007). This decline may be faster than in AD patients but slower than in PPA patients (Wicklund et al., 2007). Finally, other studies have showed that speech rate (Ash, 2019), reading performance (Irish et al., 2012), word comprehension and semantic abilities (Hardy et al., 2016; Cousins et al., 2018) do not change with time. Overall, these studies suggest that with the advancement of the disease, some speech and language impairments might become more severe in bvFTD patients. More studies are needed to understand the potential of speech and language markers in tracking bvFTD progression.

### 4.4. Current gaps and future directions

This systematic review also highlights some limitations which could guide future research in this field.

The first set of limitations concerns the samples included in the reviewed studies. Our review reveals that 80% of the included studies involved less than bvFTD 30 patients (Figure 3), which may lead to a lack of power to detect subtle language impairments. Furthermore, during the selection process, many studies (163) have been excluded because they did not include a comparison group of healthy controls. Finally, most of the articles included came from North America, Western Europe or Australia and included English-speaking participants (Table 1). Therefore, the results of this review may not be generalizable to other underrepresented populations, languages, and cultures. Together, these limitations call for studies involving a greater number of bvFTD patients and compared to a matched group of healthy participants. Multi-center studies should also be led worldwide, including currently underrepresented regions such as Africa, Asia, Eastern Europe or South America.

The second set of limitations is related to the speech and language assessments. Although a large number of studies were included in the present review, most of them investigated language as a secondary outcome measure using quick assessment tools. Therefore, the results were rarely interpreted or discussed in the original papers. Also, some domains such as motor speech, phonology and prosody have been scarcely studied until now (Figure 2), although some of them show potential as clinical markers of bvFTD. Furthermore, the assessment tools used in the literature are highly heterogeneous across studies, making it impossible to use meta-analytic statistical approaches. Because a complete speech and language assessment can be lengthy, connected speech approaches are gaining in popularity because of their high cost-effectiveness. Indeed, these approaches are quick, they allow for an assessment of multiple speech and language subdomains simultaneously and they are becoming more automated. Previous studies have shown their utility in PPA to distinguish the 3 subtypes and have studied neural correlates of connected speech components (Wilson et al., 2010a). Once further validated, they will be highly helpful in the characterization of bvFTD patients. In our review, we showed that connected speech seem to be sensitive to impairments in most speech and language domains, except for grammar.

Finally, more studies investigating the heterogeneity of speech and language symptoms in bvFTD, by comparing bvFTD subgroups, are needed to confirm the preliminary conclusions proposed in this review.

## 4.6. Conclusion

Through this review, our aim was to investigate language impairments in bvFTD syndrome, yet mostly characterized by behavioral and personality changes. Our review shed light on both extensive and heterogeneous speech and language alterations in bvFTD patients, more consistent in lexico-semantic, orthography and prosody domains. It also highlights that a globally altered cognitive status, a frontotemporal predominant atrophy and the presence of a *MAPT* mutation may be key features for language impairments. A better acknowledgement of these language and speech difficulties may help a better characterization of bvFTD and it may be helpful for both differential diagnosis and follow-up.

## Data Availability

The data is available on request.

## Acknowledgements

We thank Dr. Evans Whitaker from the UCSF Health Sciences Library for his help with the literature search strategy.

AG received an internship grant from the French Society of Neurology.

AMG is supported by CONICET; ANID, FONDECYT Regular [grant numbers 1210176 and 1210195]; Programa Interdisciplinario de Investigación Experimental en Comunicación y Cognición (PIIECC), Facultad de Humanidades, USACH.

MM is supported by postdoctoral funding from Canadian Institutes of Health Research (CIHR) and Fonds Québécois de Recherche en Santé (FRQS).

## Declaration of interest

None

## Funding

This research did not receive any specific grant from funding agencies in the public, commercial, or not-for-profit sectors.

## Supplementary materials

**Appendix. Modified Newcastle-Ottawa Scale and scoring guide. http://www.ohri.ca/programs/clinical_epidemiology/oxford.asp**

### (1) Representativeness of the sample (is the case definition adequate?)

1 point: Requires some independent validation by a multidisciplinary team. The case was defined using multiple sources of information (neurology, neuropsychology, etc.)

0.5 point: No independent validation, but the case was defined using multiple sources of information (neurology, neuropsychology, etc.) 0 point: No description, not done

### (2) Sample size

1 point: Sample size was greater than 30 participants.

0.5 point: Sample size was between 20 and 30 participants.

0 point: Sample size was less than 20 participants or a convenience sample.

### (3) Comparability of cases and controls based on the design or analysis

1 point: Either cases and controls must be matched in the design or confounders (i.e., age, sex, education) must be adjusted for in the analysis.

0.5 point: Cases and controls are only partially matched in the design or confounders (i.e., age, sex, education).

0 point: Both cases and control are not matched in the design and confounders are not adjusted for in the analysis.

### (4) Ascertainment of language impairment

1 point: Validated measurement tool.

0.5 point: Both validated and non-validated measurement tool. 0 point: no description.

### (5) Quality of descriptive statistics reporting

1 point: Reported descriptive statistics to describe the population (*e*.*g*., language measures) with a proper measure of dispersion (*e*.*g*., standard deviation, standard error, effect size, range, distribution on the graph).

0 point: Descriptive statistics were not reported, were incomplete, or did not include proper measures of dispersion.

**Legend: This scale, the scoring of which ranges from 0 to 5, assesses quality in several domains: sample representativeness and size, comparability between patients and controls, ascertainment of language impairment, and statistical quality. Studies were judged to be of low risk of bias (**≥**3 points) or high risk of bias (<3 points)**.

**Supplementary table 1:**
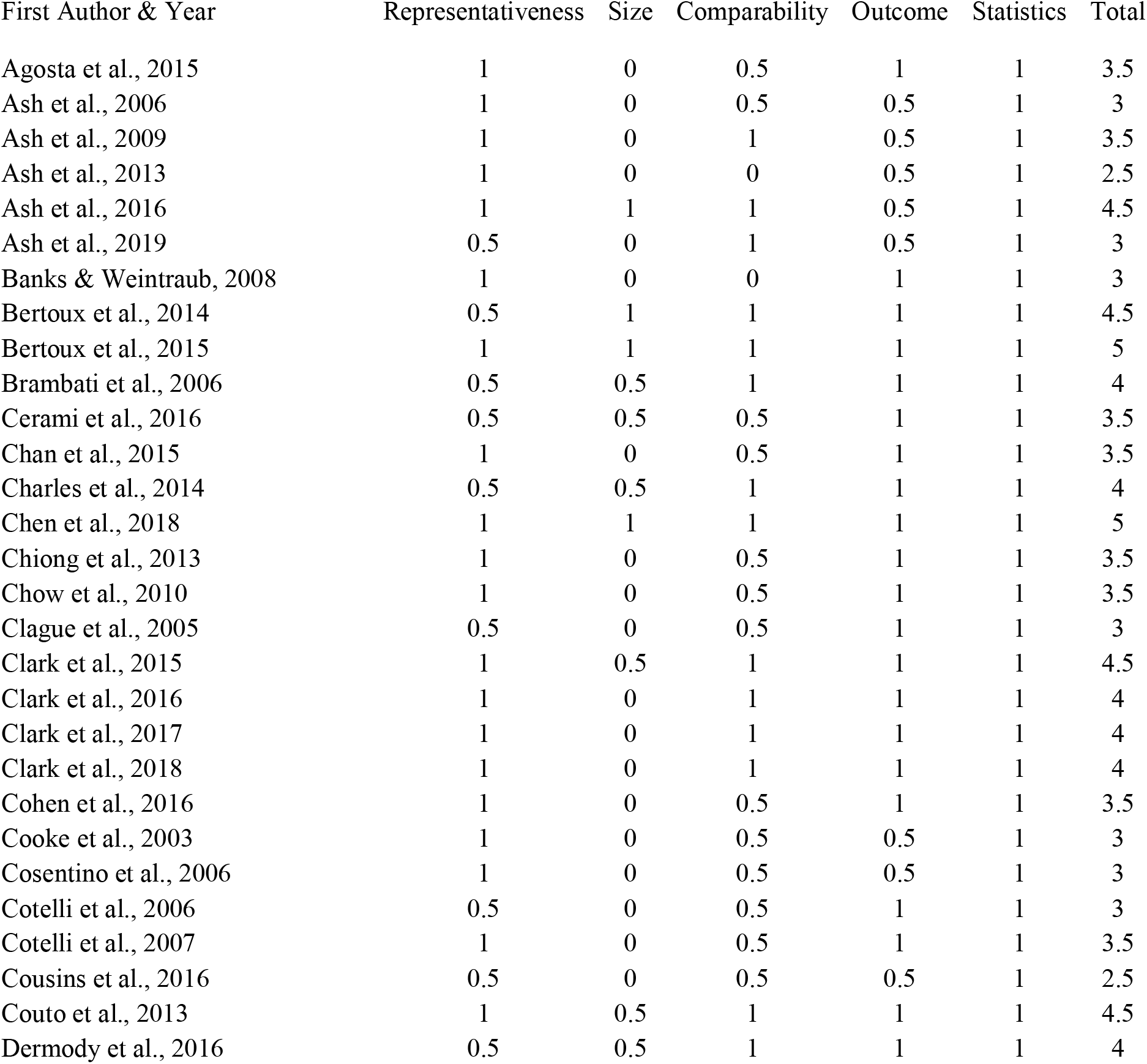
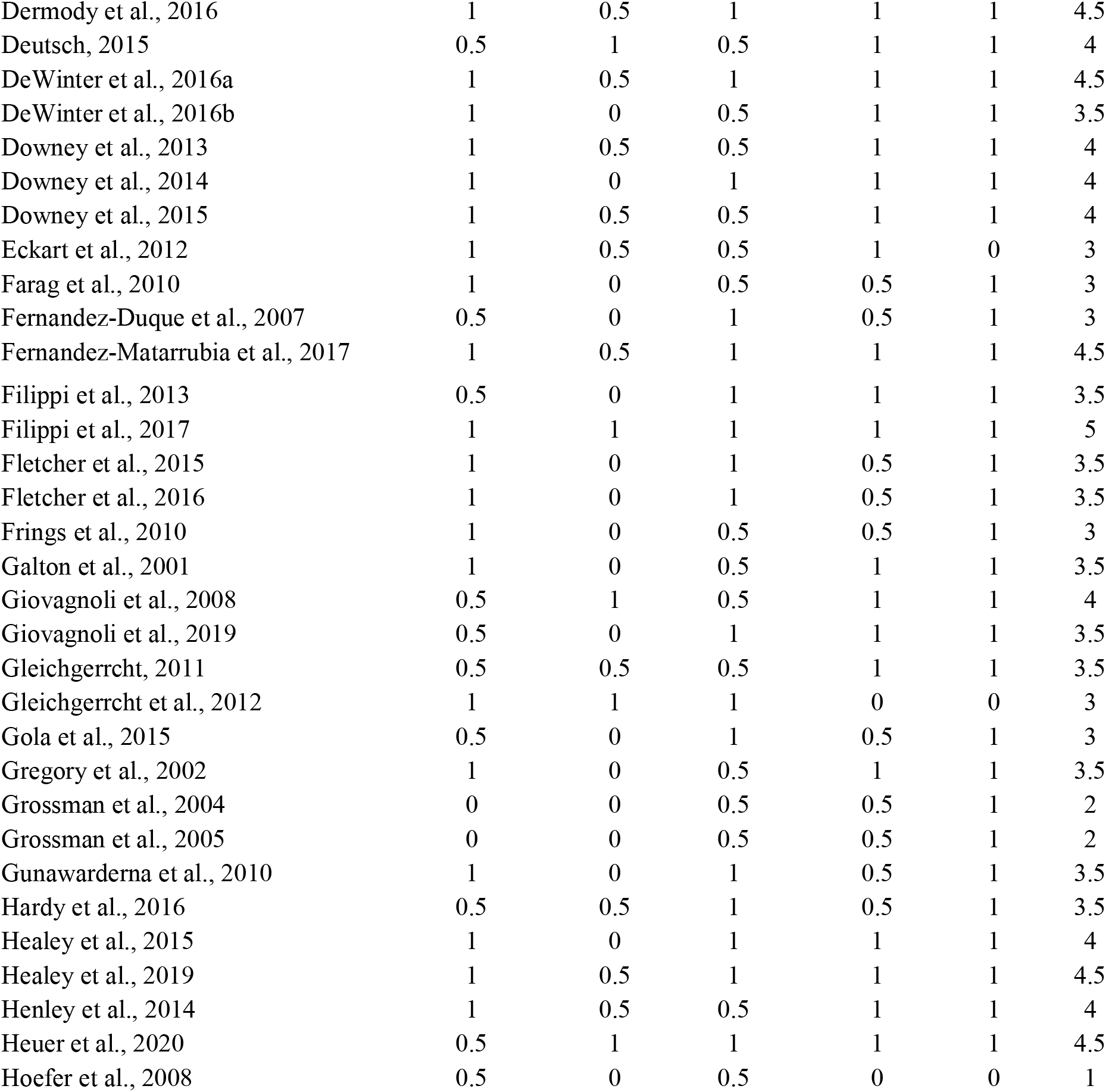
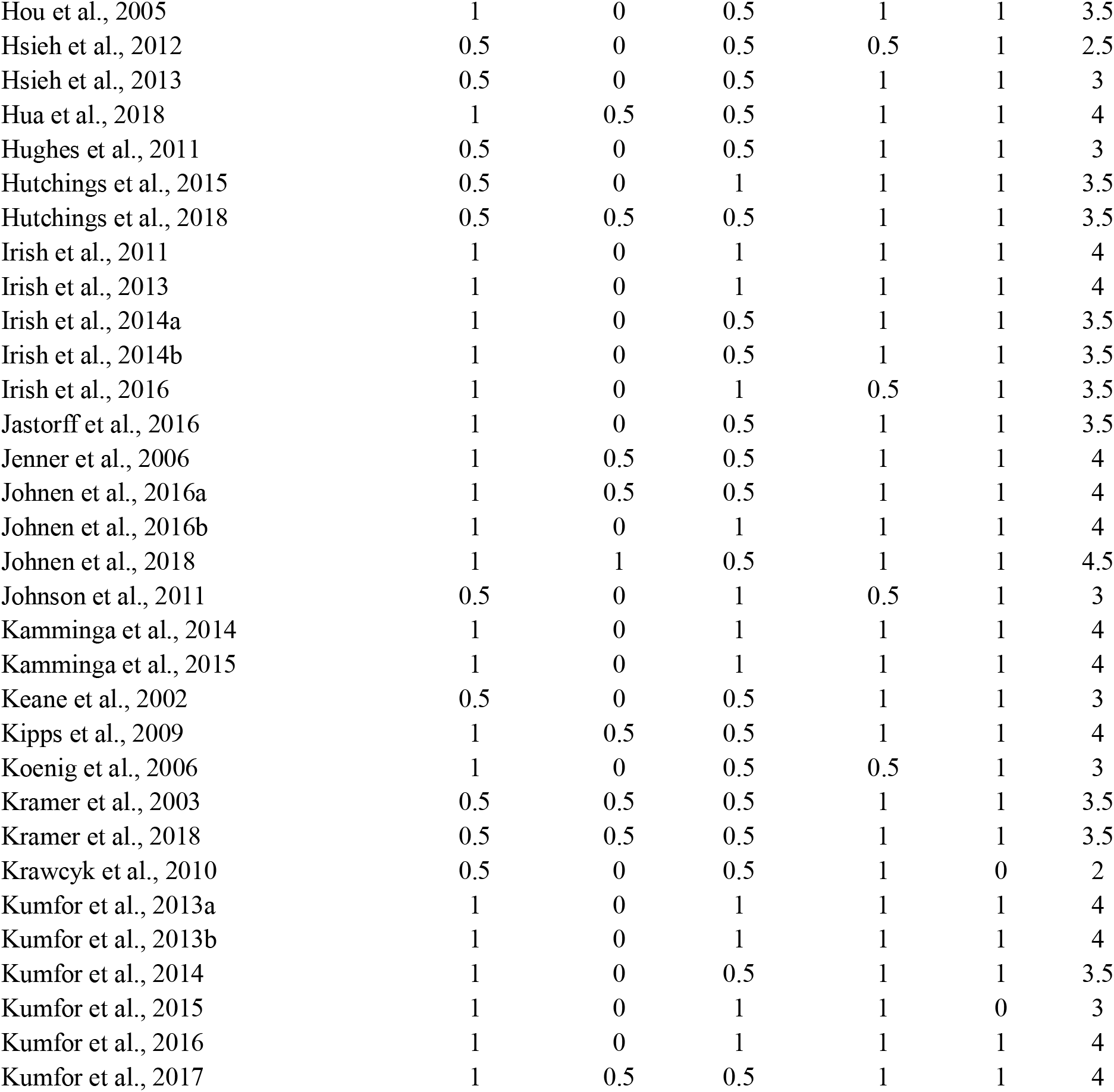
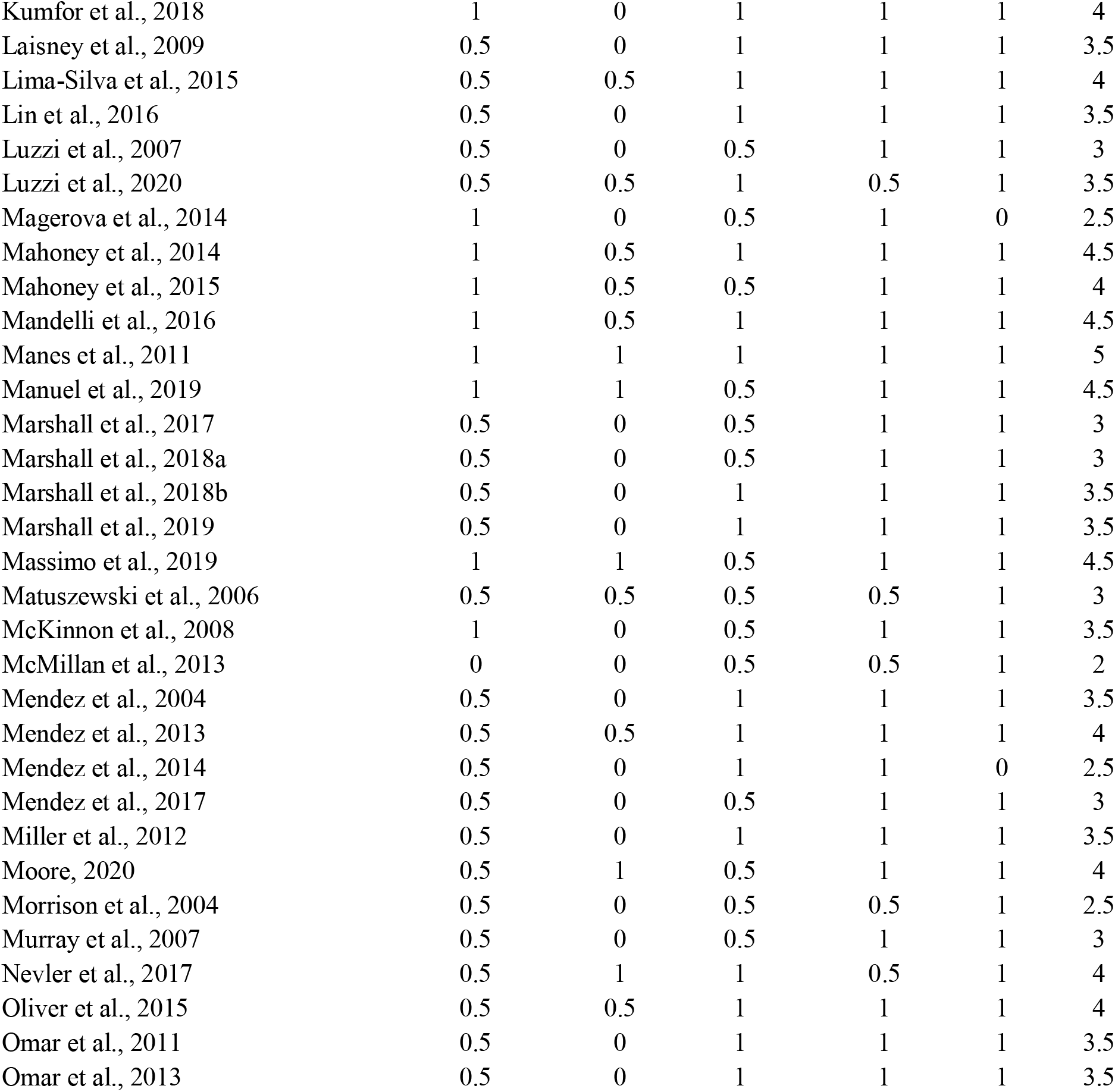
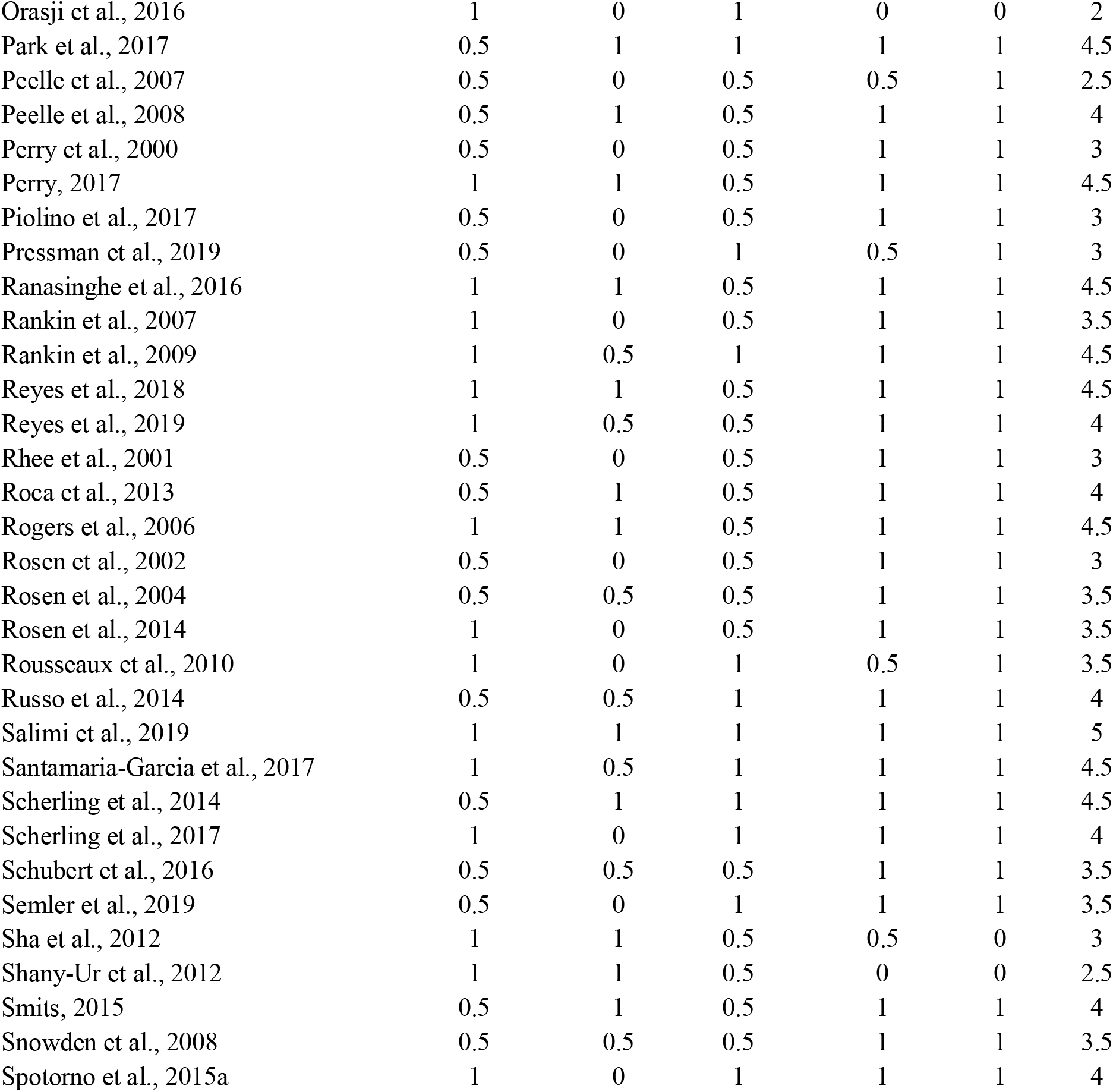
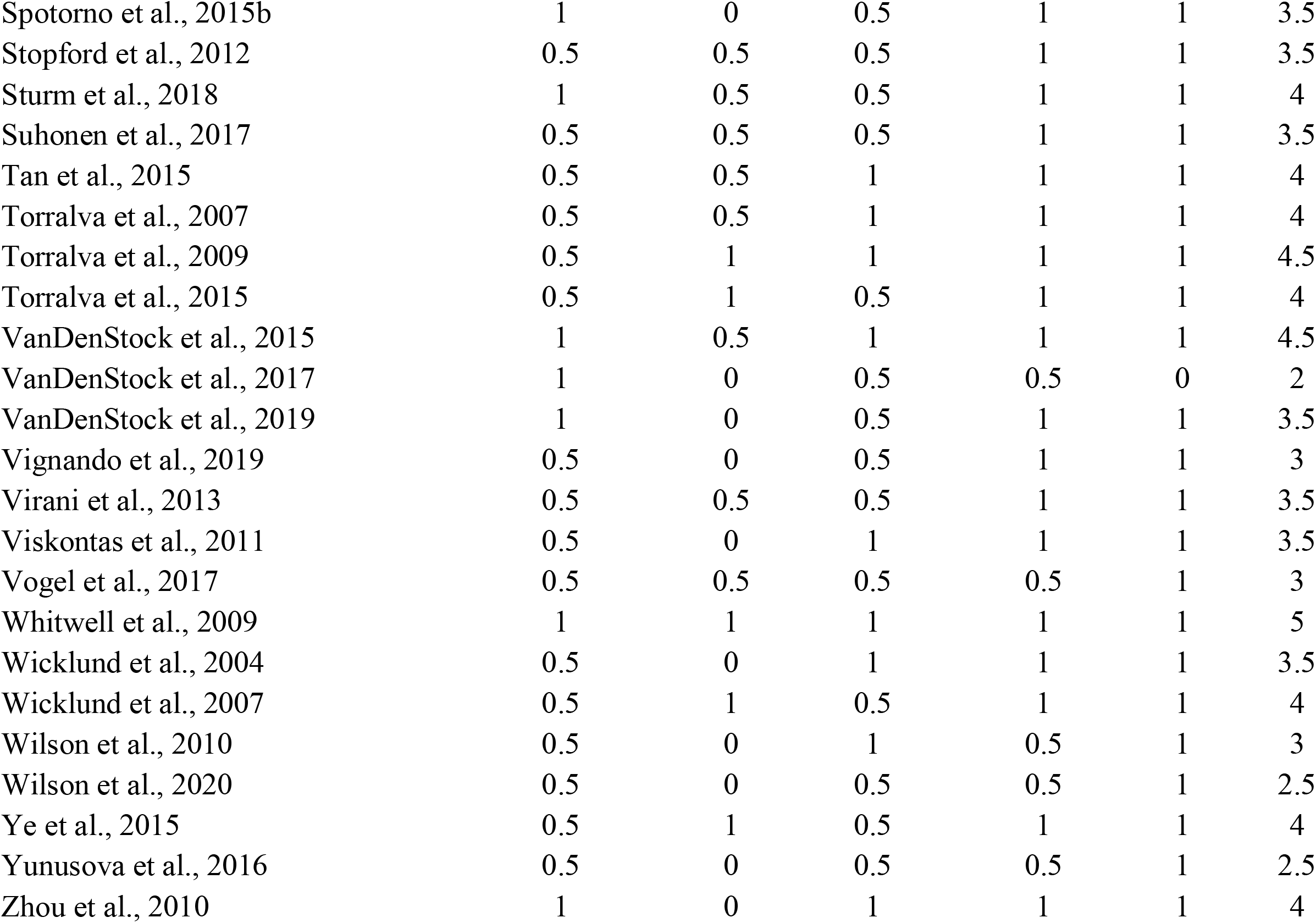
Risk of bias assessment for each study included in the systematic review.

PRISMA 2020 Checklist
| Section and Topic | Item # | Checklist item | Location where item is reported |
| --- | --- | --- | --- |
| **TITLE** |  |  |  |
| Title | 1 | Identify the report as a systematic review. | YES |
| **ABSTRACT** |  |  |  |
| Abstract | 2 | See the PRISMA 2020 for Abstracts checklist. | YES |
| **INTRODUCTION** |  |  |  |
| Rationale | 3 | Describe the rationale for the review in the context of existing knowledge. | YES |
| Objectives | 4 | Provide an explicit statement of the objective(s) or question(s) the review addresses. | YES |
| **METHODS** |  |  |  |
| Eligibility criteria | 5 | Specify the inclusion and exclusion criteria for the review and how studies were grouped for the syntheses. | YES |
| Information sources | 6 | Specify all databases, registers, websites, organisations, reference lists and other sources searched or consulted to identify studies. Specify the date when each source was last searched or consulted. | YES |
| Search strategy | 7 | Present the full search strategies for all databases, registers and websites, including any filters and limits used. | YES |
| Selection process | 8 | Specify the methods used to decide whether a study met the inclusion criteria of the review, including how many reviewers screened each record and each report retrieved, whether they worked independently, and if applicable, details of automation tools used in the process. | YES |
| Data collection process | 9 | Specify the methods used to collect data from reports, including how many reviewers collected data from each report, whether they worked independently, any processes for obtaining or confirming data from study investigators, and if applicable, details of automation tools used in the process. | YES |
| Data items | 10a | List and define all outcomes for which data were sought. Specify whether all results that were compatible with each outcome domain in each study were sought (e.g. for all measures, time points, analyses), and if not, the methods used to decide which results to collect. | YES |
|  | 10b | List and define all other variables for which data were sought (e.g. participant and intervention characteristics, funding sources). Describe any assumptions made about any missing or unclear information. | YES |
| Study risk of bias assessment | 11 | Specify the methods used to assess risk of bias in the included studies, including details of the tool(s) used, how many reviewers assessed each study and whether they worked independently, and if applicable, details of automation tools used in the process. | YES |
| Effect measures | 12 | Specify for each outcome the effect measure(s) (e.g. risk ratio, mean difference) used in the synthesis or presentation of results. | YES |
| Synthesis methods | 13a | Describe the processes used to decide which studies were eligible for each synthesis (e.g. tabulating the study intervention characteristics and comparing against the planned groups for each synthesis (item #5)). | YES |
|  | 13b | Describe any methods required to prepare the data for presentation or synthesis, such as handling of missing summary statistics, or data conversions. | YES |
|  | 13c | Describe any methods used to tabulate or visually display results of individual studies and syntheses. | YES |
|  | 13d | Describe any methods used to synthesize results and provide a rationale for the choice(s). If meta-analysis was performed, describe the model(s), method(s) to identify the presence and extent of statistical heterogeneity, and software package(s) used. | YES |
|  | 13e | Describe any methods used to explore possible causes of heterogeneity among study results (e.g. subgroup analysis, meta-regression). | YES |
|  | 13f | Describe any sensitivity analyses conducted to assess robustness of the synthesized results. | N.A. |
| Reporting bias assessment | 14 | Describe any methods used to assess risk of bias due to missing results in a synthesis (arising from reporting biases). | YES |
| Certainty | 15 | Describe any methods used to assess certainty (or confidence) in the body of evidence for an outcome. | N.A. |

PRISMA 2020 Checklist
| Section and Topic | Item # | Checklist item | Location where item is reported |
| --- | --- | --- | --- |
| assessment |  |  |  |
| **RESULTS** |  |  |  |
| Study selection | 16a | Describe the results of the search and selection process, from the number of records identified in the search to the number of studies included in the review, ideally using a flow diagram. | YES |
|  | 16b | Cite studies that might appear to meet the inclusion criteria, but which were excluded, and explain why they were excluded. | YES |
| Study characteristics | 17 | Cite each included study and present its characteristics. | YES |
| Risk of bias in studies | 18 | Present assessments of risk of bias for each included study. | YES |
| Results of individual studies | 19 | For all outcomes, present, for each study: (a) summary statistics for each group (where appropriate) and (b) an effect estimate and its precision (e.g. confidence/credible interval), ideally using structured tables or plots. | YES |
| Results of syntheses | 20a | For each synthesis, briefly summarise the characteristics and risk of bias among contributing studies. | YES |
|  | 20b | Present results of all statistical syntheses conducted. If meta-analysis was done, present for each the summary estimate and its precision (e.g. confidence/credible interval) and measures of statistical heterogeneity. If comparing groups, describe the direction of the effect. | YES |
|  | 20c | Present results of all investigations of possible causes of heterogeneity among study results. | YES |
|  | 20d | Present results of all sensitivity analyses conducted to assess the robustness of the synthesized results. | N.A. |
| Reporting biases | 21 | Present assessments of risk of bias due to missing results (arising from reporting biases) for each synthesis assessed. | YES |
| Certainty of evidence | 22 | Present assessments of certainty (or confidence) in the body of evidence for each outcome assessed. | N.A. |
| **DISCUSSION** |  |  |  |
| Discussion | 23a | Provide a general interpretation of the results in the context of other evidence. | YES |
|  | 23b | Discuss any limitations of the evidence included in the review. | YES |
|  | 23c | Discuss any limitations of the review processes used. | YES |
|  | 23d | Discuss implications of the results for practice, policy, and future research. | YES |
| **OTHER INFORMATION** |  |  |  |
| Registration and protocol | 24a | Provide registration information for the review, including register name and registration number, or state that the review was not registered. | N.A. |
|  | 24b | Indicate where the review protocol can be accessed, or state that a protocol was not prepared. | N.A. |
|  | 24c | Describe and explain any amendments to information provided at registration or in the protocol. | N.A. |
| Support | 25 | Describe sources of financial or non-financial support for the review, and the role of the funders or sponsors in the review. | YES |
| Competing interests | 26 | Declare any competing interests of review authors. | YES |
| Availability of data, code and other materials | 27 | Report which of the following are publicly available and where they can be found: template data collection forms; data extracted from included studies; data used for all analyses; analytic code; any other materials used in the review. | YES |

## Notes

### Competing Interest Statement

The authors have declared no competing interest.

